# Pandemic Control in Econ-Epi Networks

**DOI:** 10.1101/2020.08.19.20178087

**Authors:** Marina Azzimonti, Alessandra Fogli, Fabrizio Perri, Mark Ponder

## Abstract

We develop an ECON-EPI network model to evaluate policies designed to improve health and economic outcomes during a pandemic. Relative to the standard epidemiological SIR set-up, we explicitly model social contacts among individuals and allow for heterogeneity in their number and stability. In addition, we embed the network in a structural economic model describing how contacts generate economic activity. We calibrate it to the New York metro area during the 2020 COVID-19 crisis and show three main results. First, the ECON-EPI network implies patterns of infections that better match the data compared to the standard SIR. The switching during the early phase of the pandemic from unstable to stable contacts is crucial for this result. Second, the model suggests the design of smart policies that reduce infections and at the same time boost economic activity. Third, the model shows that reopening sectors characterized by numerous and unstable contacts (such as large events or schools) too early leads to fast growth of infections.

## 1 Introduction

The COVID-19 pandemic of 2020 presents a formidable challenge to policymakers: for the first time in decades they face a trade-off between epidemiological costs (lives) and economic costs (livelihoods). The key question that motivates this paper is how to design *smart* policies which are effective in reducing the spread of the disease while at the same time minimizing economic costs.

Our point of departure is that the spread of infections and economic activity happen through the same network of human interactions. We develop an ECON-EPI network model of a city, characterized by three components. The first one is the network of human interactions that specifies contacts among individuals through different network layers. The second one is the ECON component, which describes how economic activity is created on the network. The last one is the EPI component, specifying how the disease spreads through individuals across the city.

When a pandemic hits, several links of the network are severed, either as a consequence of mitigation policies or because individuals change their behavior as a response. Our key insight is that the dynamic consequences, both in terms of infections and economic outcomes, strongly depend on the type of links that are severed. Cutting certain types of links has a large impact on infections and a relatively small economic cost, while cutting other types has only a marginal impact on the infection levels but large economic costs. We find that the ECON-EPI network constitutes a useful framework to understand and measure the epidemiological and economic costs of limiting different types of social interaction, and therefore it provides guidance in designing effective policies to control the pandemic while preserving economic activity.

We start by describing a multilayered network model, of the type commonly used in the epidemiological literature. Individuals in the network differ in several dimensions (such as age, family structure and work characteristics) and interact with each other through different network layers. These layers capture the main contexts of interaction among individuals in a city, such as homes, neighborhoods, schools, public transportation, stores, entertainment venues, and workplaces. These different social contexts are also often the target of actual mitigation policies aimed at limiting the extent of interaction allowed (such as the closure of schools, the limits on large gatherings, or the shut down of non-essential businesses). Some layers (such as family and neighborhood) feature frequent and repeated interactions among a small set of individuals. Other layers (such as public transportation or shopping venues) feature less frequent and more randomized contacts among a larger set of individuals. These differences play a critical role in the speed of diffusion of the disease throughout the city, more numerous and random contacts leading to faster spreading.

Next we introduce the EPI component, which describes the dynamics of the disease. We model the progression of the disease as in the standard Susceptible-Infected-Recovered (SIR) model, while allowing for heterogeneity among infected individuals in the manifestation of symptoms and in the transmissibility upon contact. If we interpret the individuals in the economy as nodes in the network, we can express the key difference between the standard SIR set-up and our network model in terms of the infection probability of susceptible nodes. In the SIR set-up, this probability is the same for all nodes and depends on the average number of infected nodes in the system. In the network set-up, it differs across nodes, and for each node it depends on its own fraction of infected connections. This difference is crucial to understand why the SIR and the network model generate very different infection dynamics.

The last component of the network model is the ECON one, which specifies the details of production and the links between workers and shoppers. In the economy, a homogenous final good is produced in establishments by a stable team of workers and by capital. Each establishment belongs to one of two sectors: High-contact and Low-contact (H- and L-henceforth). These two sectors are a parsimonious way of classifying actual sectors into two groups depending on the strength of the link between production and spreading of the disease. In order to identify these sectors, we use information from the Occupation Information Network (ONET) database to construct measures of physical proximity and frequent interaction between workers and customers in each 2 digit NAICS sector. We then classify the actual sectors that score above the average in both measures as belonging to the H-sector. Examples of these sectors are retail, food, accommodation, and health. Production in the H-sector is likely to cause fast spreading in a pandemic for two reasons. First, H-workers cannot produce from home and thus are more exposed to the disease. Second, as they have numerous and randomly selected daily contacts with customers, they are more likely to spread the disease. The remaining sectors are classified as belonging to the L-sector. In the L-sector, production involves minimal physical proximity and/or interaction with customers. These features imply that production in the L-sector has less impact on infection spreading for two reasons. First, workers in the L-sector have fewer and more stable contacts as they only interact with other workers in their team. Second, a significant fraction of them has the ability to work from home. In addition to the difference in spreading potential, we allow the H and L sector to differ in terms of the marginal product of each worker, so that we can evaluate more accurately the effect on output of shut-down policies that target different sectors.

One important feature of the ECON component is that we explicitly model the productive role of customers by assuming that production in the H-sector requires both workers and customers. While workers in the H-sector are only a subset of the population, customers are drawn from the entire population, as every individual in the city shops. This implies that when a pandemic hits and most people severely limit their interactions (either because of shut-down policies, quarantine, or fear) there are two effects on production. The first one is the direct supply effect that reduces production because workers cannot work. The second one is the indirect demand effect, that reduces output because customers do not show up at H-sector establishments where they are an essential factor of production.

We next define a pre-pandemic city equilibrium where heterogeneous workers are allocated efficiently across different establishments and where the shopping capacity of the H-sector satisfies the demand of shopping trips by the population. Using restrictions from the prepandemic equilibrium together with sector level data from the New York-Newark-Jersey City (NY-NJ-PA) metro area, we can pick values for the parameters that fully characterize the ECON-component. Before we can use our set-up to conduct policy experiments, we need to pin down two additional set of parameters. The first is the number of contacts between different types of nodes before and during the pandemic. The second is the set of parameters that governs disease progression and transmission (the EPI component). We use the seminal work of Mossong et al. (2008) to pin down the amount of contacts in the pre-pandemic equilibrium, and data from Google Mobility reports to capture the reduction in contacts during the pandemic. Regarding the EPI component, we calibrate its parameters using both evidence from epidemiological studies and by matching key moments of the infection’s early phase in the NY-NJ-PA metro area.

Finally, we use the fully calibrated model to perform a number of experiments. The first experiment compares the dynamics of infection in our set-up with the standard SIR set-up. We put the two models on equal footing by choosing parameters in both models to match the dynamics during the early phase (March 8th - April 3rd) of the pandemic in the NY-NJ-PA metro area. We then compare their predictions for the second phase of the pandemic (April 3rd - April 26th) with the data. The main result is that the standard SIR model implies a counterfactually fast spreading of the infection, while the network set-up predicts a plateau of infections, as observed in the data. The reason for this difference is that in the SIR set-up new infections depend on the average fraction of infected nodes in the system so, once total infections reach a critical level (as they did in New York), it progresses rapidly until herd immunity is reached. In the network model, however, infections depend on the number of local contacts. Therefore, it is possible that some areas of the network remain untouched by the infection, while at the same time the disease dies out in other areas due to herd immunity at a “local” level.

Our second experiment uses the ECON-component of the model more intensively to study “smart” mitigation policies that can achieve better outcomes during the lock-down phase. These policies can be valuable during a possible second wave of COVID-19, or in future pandemics. We find that policies that reduce the workers shutdown in the L-sector, while at same time increase the workers shutdown in the H-sector can, under some circumstances, achieve a double gain; that is, reduce the spreading of the disease and simultaneously reduce the output loss. It is immediate to see that such policies the spread of the infection. The outcome for output depends on the relative marginal product of labor in the two sectors, which in turn is a function of the amount of capital and of the intensity of the shutdown in each sector. We show that in our NY-NJ-PA metro area case study, for the observed level of shutdown and for the calibrated level of capital in the two sectors, a policy involving a substantial double gain (reduction in infection cases equal to 1.5% of the population and 1% increase in output) could have been implemented.

Our third and final set of experiments concerns the reopening of the business sectors and schools, once the pandemic has passed its peak. We find that the timing and extent of the reopening are crucial. A broad reopening, which includes the H sector or the schools, at a time when the level of infections is still significant in the city inevitably leads to a large second wave. Our set-up suggests two reopening strategies that could avoid a second wave. The first one is to prolong a wide-spread shut down and then reopening only when the level of infections is minimal. The second one is to start the reopening early, but only in the L-sector, which can achieve substantial output gains with little infection growth.

There are three important lessons that we learn from our work. First, the micro structure of the network is essential to understand and predict aggregate infection dynamics. When connections are random and unstable across the network (like in the standard SIR model), an infectious disease spreads fast. When connections are instead clustered and repeated, the same disease stays local and dies out. Layers in our ECON-EPI network lie in between these two extremes, with some layers (the H-sector) being random and unstable and some others (like the family) being more clustered and stable. The dynamics of infection in a city depends on the relative importance of these layers, and policies geared to contain infections are most effective when they can target different layers separately.

Second, in order to assess the economic cost of policies aimed at containing the infection it is important to specify the micro structure of production. The cost of shutting down a worker is its marginal product. In an undistorted equilibrium, marginal product is captured by the wage; however, during widespread shutdowns like those observed during the COVID-19 pandemic, marginal product can be different (and higher) than the wage, and thus the cost of alternative shutdown policies can be assessed only by specifying the details of production.

Finally, our set-up suggests that there are important complementarities between various types of mitigation policies. For example, we find that reopening schools is only viable if it is preceded by a strict lock-down of many economic activities, which brings infections to a minimal level. Also, we find that when people adopt practices that reduce the transmissibility of the disease (e.g. wearing face-masks), policies that reduce contacts are more effective. A key insight is that the use of a structural model of interaction is necessary to understand and quantify the extent of these complementarities.

The paper is structured as follows. Section 2 summarizes the related literature. Sections 3 and 4 describe the ECON-EPI network and our calibration strategy. Section 5 shows how the network model can help explain the data. Section 6 discusses the policy experiments and Section 7 concludes.

## 2 Connection to existing literature

The COVID-19 pandemic of 2020 has spurred a new and fast growing literature at the interface between epidemiology and economics, studying the effects, both on infection and economic outcomes, of different policies geared to containing the spreading of the disease.

A first generation of papers has modeled the epidemiological component using versions of the standard SIR random mixing model, as in Kermack and McKendrick (1927). Examples of these works include Acemoglu et al. (2020), Alvarez et al. (2020), Atkeson (2020), Eichenbaum et al. (2020), Favero et al. (2020), Glover et al. (2020) and Jones et al. (2020).

Modern research in epidemiology has moved beyond this classical framework to explicitly model the patterns of interaction among agents and makes extensive use of network theory to predict the pattern of infections in a city or in a country.^1^ One of the main contributions of our paper is to integrate the network modeling of infection from epidemiology in an economic model of a city, where the network plays an explicit role both in the transmission of infection and in the creation of economic value. We now briefly discuss how our paper is different from other recent and excellent works that also use network theory to study the COVID-19 pandemic. Karaivanov (2020) analyzes the diffusion of COVID-19 in an abstract network and makes the point that transmission is different from the one in the standard SIR model. However he restricts his attention to the epidemiological component. Baqaee et al. (2020) and Akbarpour et al. (2020) both use a network framework to analyze the economic and epidemiological effects of containment and re-opening policies. Baqaee et al. (2020) focus on aggregate (US) outcomes, while Akbarpour et al. (2020) focus, as we do, on metro-level outcomes. There are two important differences that distinguish our work from theirs. The first is that we model the network differently. In their works the main heterogeneity across nodes rests on the number of contacts. Nodes in our network, in addition to being heterogenous in terms of number of contacts, are also heterogenous in terms of the pool from which they draw their daily contacts.^2^ This feature of the contacts, which we refer to as “stability”, not only is empirically relevant, as it captures the different degree of randomness of daily contacts in different occupations, but is also quantitatively important to explain infection dynamics. The second difference is in the modeling of the production structure. Both papers assume labor is the only factor of production, while we use an establishment production function that, in addition to labor, uses capital and (for retail establishments) customers as inputs. This production function allows to evaluate the output costs of workers’ shutdown more accurately, as well as the impact on output of the reduction in shopping contacts (i.e. demand effects).

Our work is also related to a number of more empirical studies exploring the role of heterogeneity across sectors and across workers in the spreading of the infection and in designing efficient containment policies, such as Benzell et al. (2020), Dingel and Neiman (2020), Kaplan et al. (2020), Leibovici et al. (2020), and Mongey et al. (2020).

## 3 The ECON-EPI network

We now describe the details of the ECON-EPI network, a model designed to capture human and economic interaction in a typical US metropolitan area. We first present the network structure, i.e. the links that connect individuals in their different activities. We then proceed to specify the EPI component, i.e. how infections progress and spread through the network.

Finally, we describe the ECON component, i.e. how interactions in the network produce output. In this part, we first specify a pre-pandemic steady state equilibrium, which describes the normal state of economic affairs before the pandemic. We then discuss how the arrival of a disease and the adoption of containment measures affect economic activity during the pandemic period.

### 3.1 A multilayered network

We construct a multilayered network where individuals of different characteristics (age, employment status, public transportation usage, etc.) interact with each other. The set-up is necessarily stylized. Nevertheless, it has enough richness to capture key aspects of the social distancing policies that have been implemented during the 2020 COVID-19 pandemic. Time is discrete and the network is generically represented by a *M × M*, time varying, adjacency matrix G_t_, where each node represents an individual. Individuals are heterogeneous in several dimensions. In terms of age, there are adults and kids. Kids are a fraction *ν_K_* of total population and go to schools. In terms of employment characteristics, adults may work in different sectors or be out of the workforce. Additionally, individuals differ in the size of their household, their number of neighbors, and their use of public transportation. We now proceed to describe the various layers connecting individuals. These layers affect the probability for each individual of contracting and spreading the disease throughout the network.

#### Households and Neighbors

Households can be single member (composed of one adult) or two-member, composed of an adult and a kid. Members of the same household are fully connected through intra-household links. These links form the first layer of our network, contained in the adjacency matrix G^*H*^. The left panel of Figure 1 shows an example of household links in a city with 12 households, where the circles represent adult members and the stars represent kids. Households are placed next to each other on a ring (as in Watts and Strogatz 1998), and each household member is connected all the members of *N* neighboring households on the left and on the right. The neighborhood links form the second layer of our network and are recorded in the adjacency matrix G^*N*^. Household and neighborhood links are ‘short-stable links,’ meaning that they are active at every point in time, and connect individuals who are close to each other.

**Figure 1:**
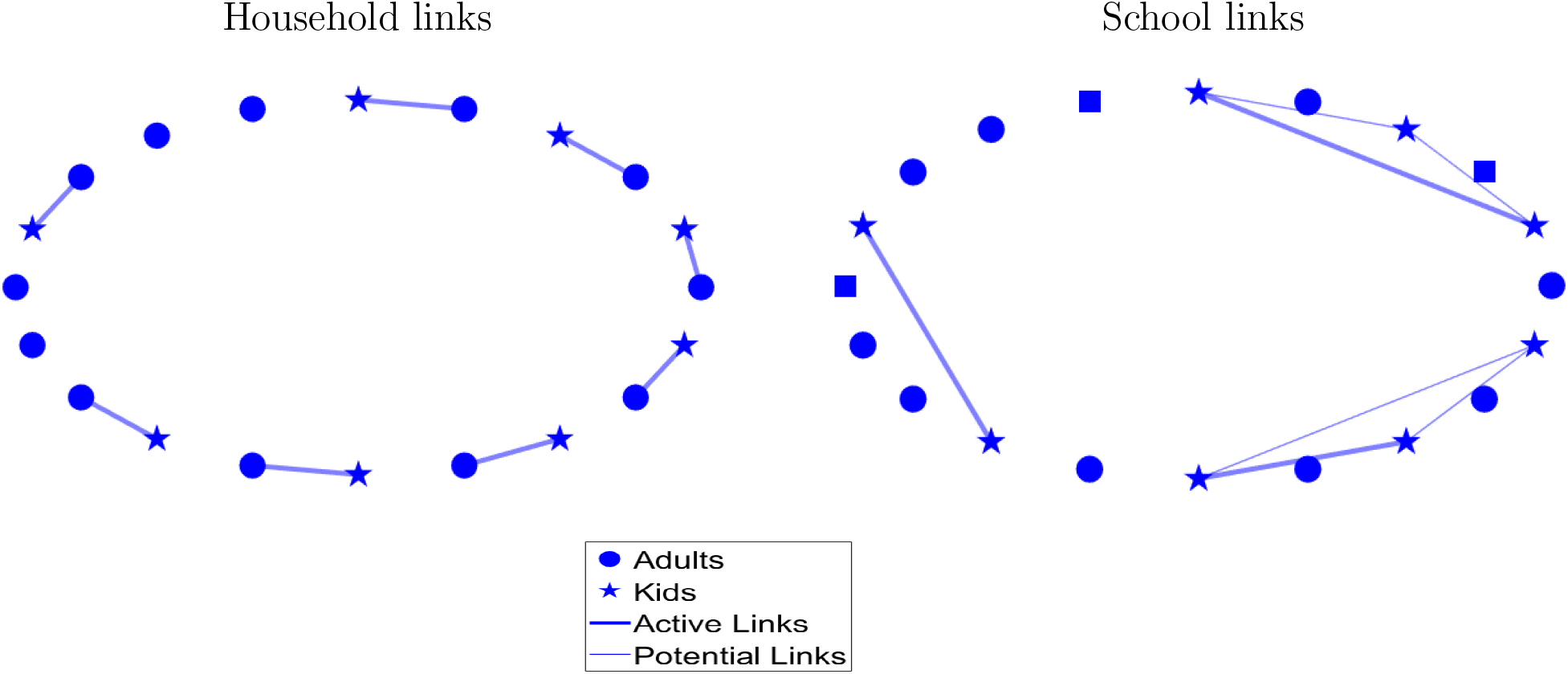
Households and Schools

#### Schools

Our third layer involves a school system where each day every kid interacts with a subset of other kids in her school. Each school draws kids which live close to each other, and the school size *Q* determines the pool of potential interactions of each kid. We refer to these links as “potential” school links and their associated adjacency matrix is denoted by G^*S*^. The right panel of Figure 1 shows the potential and active links for three schools (two of size 3 and one of size 2) for the same example network in the left panel of the figure. Note that we refer to school links as “potential” because, in contrast to the links in the first two layers, they are not always active: each kid has active links only with a subset of her schoolmates, which is randomly drawn every period. We define school links as ‘short-unstable’ meaning that they connect individuals who are geographically close to each other, and that they change their status (from active to inactive) over time. The reason for introducing this layer is to later evaluate the effect of school closures and reopenings on the spreading of the infection.

#### Public Transportation

The next layer of the network specifies interactions through public transportation. A fraction *ϕ* of individuals uses public transportation. Each public transportation vehicle has a capacity of seating *P* individuals, and we assume that agents living close to each other use the same public transportation vehicle. This implies that each individual using public transportation is potentially connected to locally close individuals who also use public transportation. Potential public transportation links are summarized in the adjacency matrix G^*P*^. During each public transportation trip, each individual interacts with a random subset of the vehicle occupants. Therefore, individuals who use public transportation will be more exposed to the disease than those who use private means of transportation.^3^. Like school links, public transportation links are short and unstable. The difference between the two is that school links involve only kids, while public transportation connects adults as well as kids.

#### Workplace

A fraction of adults in the network work. The workplace layer describes how working adults interact with each other and with the rest of society. The city features two distinct workplaces, which we label L (for Low-contact) and H (for High-contact). In the L-workplace (which is meant to capture sectors like manufacturing) there are stable teams of L-workers. In the H workplace (which is meant to capture sectors like retail or hospitality) there are similar teams of workers, but these workers are also connected with a time-varying subset of customers. We now describe in more detail the two workplaces.

#### L-Workplace

L-Workers are a share *ν_L_* of adults. Some of them (e.g. software developers) have the opportunity of working from home, which they will use in different intensity before and after the pandemic. The remaining members (e.g. assembly line workers) cannot work from home, and they are all connected to each other when working. The lightly colored nodes in the left panel of Figure 2 are a team of L-workers. Note that three of them are connected to each other, while one (labeled home worker) is not connected. For production purposes the home worker is part of the team, but it can perform work without contact (and hence without risk of contagion) with the other team members.

**Figure 2:**
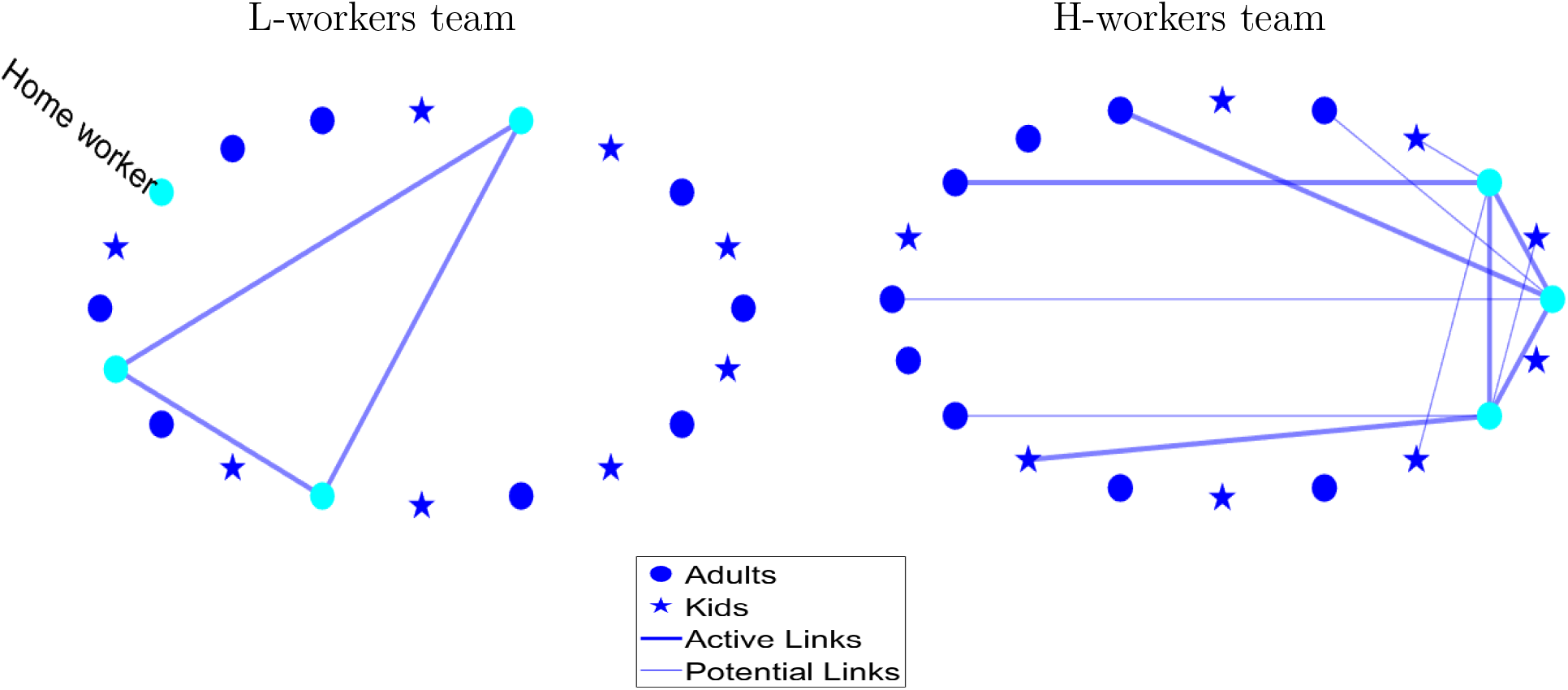
Workplaces

#### H-Workplace

H-Workers are a share *ν_H_* of adults and they represent occupations that, for the purpose of production, involve stable contacts with co-workers (just like the L-workers) as well as unstable contacts with external customers (such as retail). The right panel of Figure 2 illustrates a team of H-workers. The lightly colored nodes represent the members of the team. Note that each worker in the team has potential links with other nodes in the network (potential customers, connected to the workers by the thin lines), and in each period some of these links become active (actual customers, connected to the workers by the thick lines). Note that customers are not connected to each other. This captures the fact that individuals from certain professions (doctors, bartenders, shop clerks) may come into contact with several clients during a day, sequentially, so their visits do not overlap. Finally observe that due to the nature of their work, H-workers do not have the opportunity of working from home.

Worker links are, thus, intrinsically different from school links or public transportation links for two reasons. First, they include long links, connecting individuals in the network who are not necessarily in the same geographic area. Second, the number of connections of each individual worker can be different, with L-workers having only stable links, and H-workers having stable and unstable links. For convenience, we record worker links in two adjacency matrices: G^*W*^, which records all co-worker links (stable) in the H and L sector, and G^*C*^ which records worker to customer links in the H-sector (unstable).

#### Network Clocks

An important network feature, for the purpose of disease spreading, is the presence of unstable links between nodes. Connections in the household layer, the neighborhood layer, and among teams of workers—in the workplace layer—are stable, as individuals are linked with the same set of people every period (e.g. their network links are always active). On the other hand, interactions in the school layer, the public transportation layer, and between shoppers and workers—in the workplace layer—are inherently unstable (e.g. only a subset of potential links are active every period). To model this, we incorporate a *clock* in the spirit of Acemoglu et al. (2010) and Acemoglu et al. (2013). More specifically, for all *t ≥* 1, we associate a *clock* to every link of the form (*i,j*) in the original adjacency matrix G^*i*^(where *i* = *S, P, C*) to determine whether the link is activated or not at time *t*. The ticking of all clocks at any time is dictated by i.i.d. samples from a Bernoulli Distribution with fixed parameter *ϱ_i_ ∈* (0, 1], meaning that if the (*i,j*)-clock ticks at time *t* (realization 1 in the Bernoulli draw), the connection between agents *i* and *j* is active at time *t*. This is meant to capture two kids in the same school having lunch together on a given day, two persons sitting next to each other in the subway, or a customer and a cashier interacting over a transaction. The Bernoulli draws are represented by the *M × M* matrix of zeros and ones *c^i^_t_*. Thus, the adjacency matrices for school, public transportation and worker-customers networks evolve stochastically across time according to

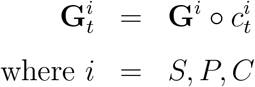

##### City Network

Finally, we superimpose the layers described so far to construct a meta network which corresponds to our synthetic city. The adjacency matrix capturing all links within a city, G_t_, is constructed as a weighted sum of the different layers. The weights correspond to the relative importance of each layer, capturing that individuals spend different amounts of time interacting with others in different social spheres. In particular, we have that

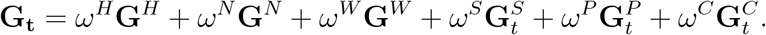

Each element in G_t_, denoted by *g_i,j,t_*, summarizes the link between two individuals *i* and *j* at time *t*, weighted by the strength of their relationship.

##### 3.2 The EPI component

The spread of the disease within our multilayered network is the result of two types of events: the person-to-person transmission of the disease (which depends on the network) and the progression of the disease for a given infected person, which is independent from the network structure. Our modeling of the disease progression closely follows a SEIR structure, a variant of the SIR model that is common in the epidemiological literature, where we added the possibility of an “asymptomatic” branch. This assumption is motivated by the fact that, during the COVID-19 pandemic, many infection cases went undetected, either because symptoms were mild, or because testing was not available. These cases were never officially recorded as infected, and transited directly to the recovered stage. However, according to several studies, they significantly contributed to the spread of the disease.^4^

Each individual node can be, at each point in time, in one of six health states: Susceptible, Exposed, Infected-Asymptomatic, Infected-Pre-symptomatic, Infected-Symptomatic, and Recovered.

1. Susceptible (S): a node which has not been exposed to the disease, but may contract it in the future.
2. Exposed (E): a node which has been in contact with an infected node and has contracted the disease. Exposed nodes are not infectious and continue to perform normal activities. However they will transit with certainty to one of the infectious states the day following the exposure.
3. Infected Pre-symptomatic (IP): a node which is infected and will show symptoms in the future. Nodes at this stage do not know they are infected, so they continue to perform normal activities. They transmit the disease with probability *π*.
4. Infected Symptomatic (IS): a node which is infected and shows symptoms. IS nodes are removed from all layers of the network, with the exception of the household layer. They transmit the disease with probability *π*.
5. Infected Asymptomatic (IA): a node which is infected, but does not and will not show severe symptoms. These nodes do not know they are infected, so they continue to perform normal activities. IA nodes, when in contact with an S node, transmit the disease with probability *ηπ*, with 0 *≤ η ≤* 1.
6. Recovered (R): a node which is no longer infected. Recovered nodes are immune to the disease and can resume normal activities.

Note that all nodes in an infected state can transmit the disease to susceptible nodes, although the infected asymptomatic are less likely to transmit. The transition between states is illustrated in Figure 3. A susceptible node *i* contracts the disease at time *t* with probability *p_i,t_* and if it does so, moves to the exposed state. An exposed node transitions to the asymptomatic stage with probability *α* and to the pre-symptomatic stage with probability 1 *− α*. A pre-symptomatic node moves to the symptomatic stage in each period with probability *γ* and a symptomatic node moves to the recovered stage with probability *ρ_S_*. An asymptomatic node, on the other hand, has a probability *ρ_A_* in each period of moving directly to the recovered stage. Finally, recovery is an absorbing state. The key object of our analysis is *p_i,t_*, the probability that a susceptible node *i* contracts the disease in period *t*. The probability *p_i,t_* is a function of the active contacts of node *i* at time *t* (encoded in G_t_), of their health status and on the odds of contracting the disease conditional on meeting an infected node (governed by the parameters *π* and *η*). In particular we can write

**Figure 3:**
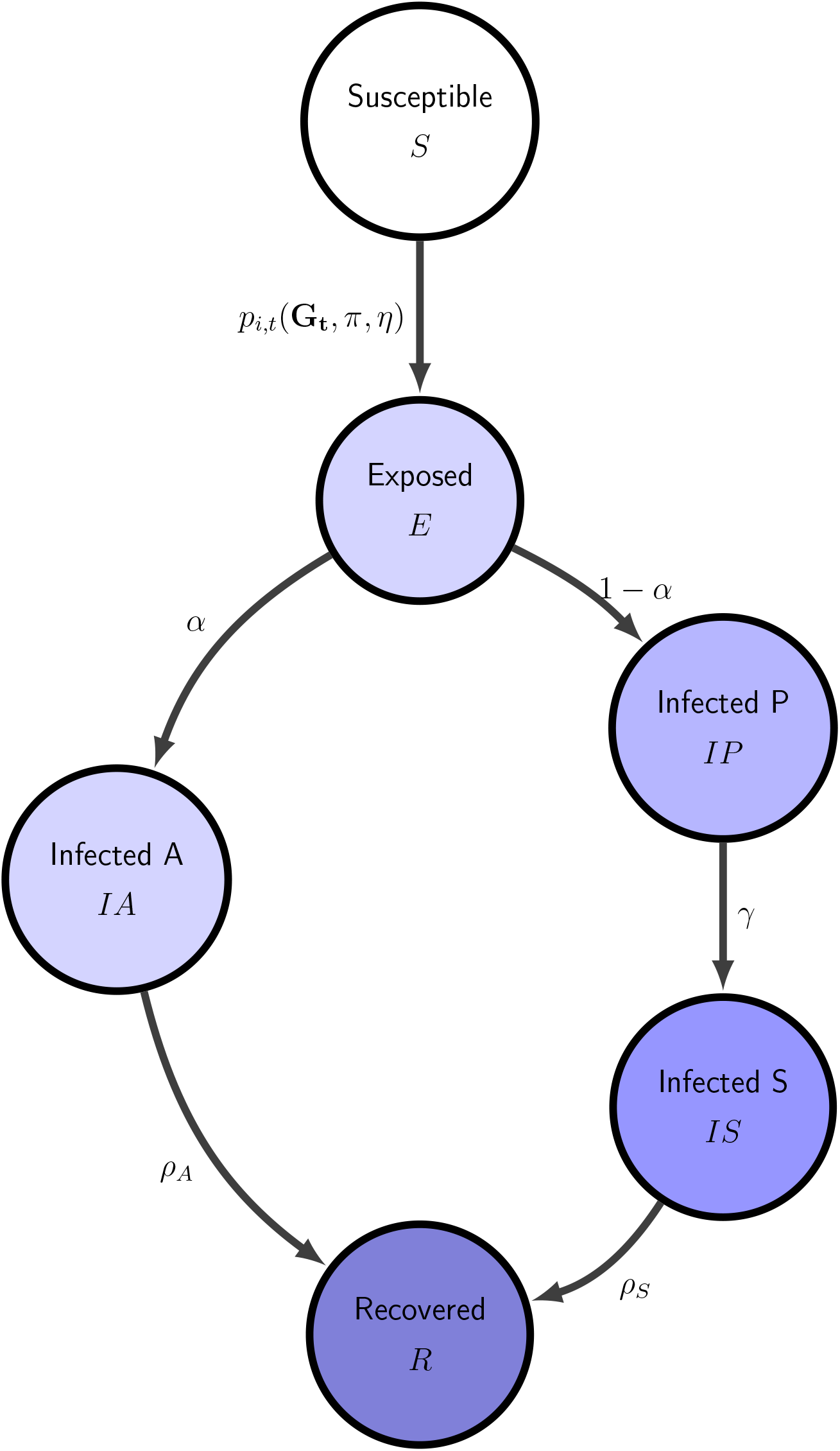
Transition between health states

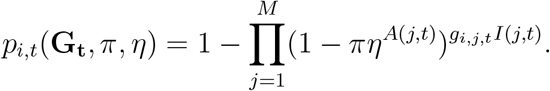

where *g_i,j,t_* is the *ith, jth* element of G_t_, *I*(*j, t*) is an indicator function that equals 1 when node *j* is infected (either pre-symptomatic, a-symptomatic or symptomatic) at time *t*, and zero otherwise, and *A*(*j, t*) is a similar indicator function for the infected-asymptomatic status. This equation makes it clear that the spreading of the disease in the economy depends not only on the disease prevalence (captured by *I*(*j, t*) and *A*(*j, t*)) and on the biological transmissibility (captured by *π* and *η*), but also on the network structure summarized by G_t_.

##### 3.3 The ECON Component

Individual nodes, together with the network structure, produce, at each point in time, new infections and economic output. This section describes how output gets produced over the network and how it is affected by social distance policies and by behavioral changes that result from the progression of the infection. The two workplaces described in Section 3.1 map into two sectors where output is produced. Both sectors produce the same homogenous good (which is also the numeraire) and production is organized in establishments. In the L-sector there are *Q_L_* ex-ante identical establishments, each endowed with the same amount of fixed capital *K_L_*. These establishments employ teams of L-workers. In the H-sector there are also establishments which hire teams of H-workers, and we allow capital to be potentially different across establishments. As we think of these two sectors as having a substantially different occupational mix, we assume that workers cannot move across sectors.^5^ In the L-sector, production requires L-workers and capital, while in the H-sector, production requires H-workers, capital, and customers. We first describe a pre-pandemic steady state equilibrium, where there are no infected nodes and the level of economic activity is stable over time, and then move on to describe how economic activity evolves as the disease hits the city and containment measures are adopted.

###### L-Sector

Recall that establishments in this sector are homogenous. Each establishment produces *y_L_* units of output according to

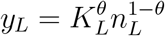

where *n_L_* denotes units of labor. Notice that *n_L_* is labor input which is not necessarily the same as employment, as not all L-workers supply the same amount of labor input. In particular, consistently with recent empirical work by Dingel and Neiman (2020) and Leibovici et al. (2020), we assume that a fraction *ω* of L-workers can work from home, and the labor input (or productivity) of these workers is *δ_ω_*% higher than the labor input of those that cannot work from home. Given the wage rate per unit of L-work *w_L_*, the establishment manager chooses labor input to maximize profits, which are given by *y_L_−n_L_w_L_*. This implies that per establishment labor demand is given by

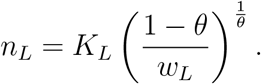

Labor supply of the L-workers is inelastic and is simply given by the total numbers of L-workers times their average labor input. A pre-pandemic equilibrium is then a wage rate *w_L_* and quantity of L-labor per plant *n_L_* such that, i) given wages, *n_L_* is chosen optimally by the plant manager and ii) labor market for L-workers clear. Equation 5 summarizes these two conditions

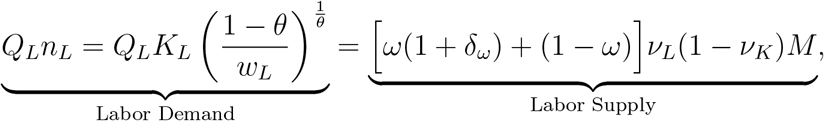

where *ν_L_* denotes the share of adults which work in the L-sector and (1 *− ν_K_*) the share of adults in the population, implying that *ν_L_*(1 *− ν_K_*)*M* is the total number of individuals who work in the sector, while *ω*(1 + *δ_ω_*) + (1 *− ω*) is their average effective labor.

###### H-Sector

The locking down of retail establishments has been at the centerpiece of the policy discussion during the 2020 COVID-19 pandemic. Although it has been widely acknowledged that larger retail establishments lead to fast spreading of the disease, there has been much less emphasis on the fact that large retail establishments are, on average, more productive (see, for example, Foster et al. 2006), and thus shutting down workers in those establishment might be more costly. In order to capture this trade-off we introduce heterogeneity in H-establishments. We consider two types of establishments: small and large, indexed by *j* = 1, 2. There are *Q_H_*_1_ small establishments (mom and pop corner stores) which have less capital, and have customer and employee bases drawn from individuals in a geographically close area. There are *Q_H_*_2_ large establishments (e.g. large shopping malls, concert venues, and stadiums) which have more capital and have customer and employee bases drawn from the entire network. Each establishment of type *j* in the H-sector produces *y_Hj_* units of output according to

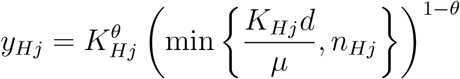

where *K_Hj_* denotes the capital of establishment of type *j* (*K_H_*_1_ *< K_H_*_2_), *n_Hj_* denotes the number of workers (which in this sector are homogenous) employed by establishment of type *j, µ* is the number of customers that a H-worker can attend to and *d* represents the number of customers (per unit of capital) which shows up at establishment *i*. This assumption captures that in the H-sector customers and workers are complement in production: if a customer does not go to the establishment, a sale does not materialize and output is not produced. In addition, if there are too few workers, they may not be able to serve all the customers that come to the establishment. The establishment manager takes as given the wage rate *w_H_* and the demand *d* and hires workers to maximize profits, which are given by *y_Hj_ − n_Hj_w_H_*. This implies that labor demand in establishment of type *j* is given by

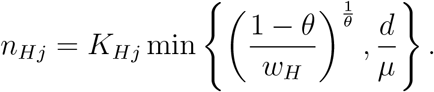

Similarly to the L-sector, the labor supply of the H-workers is inelastic and is given by the total numbers of H-workers, which is equal to *M*(1*−ν_K_*)*ν_H_*. The last element that is needed to define a pre-pandemic equilibrium is the determination of *d*. Recall that in our model city there are *M* individuals, and each person makes *s* shopping trip every period. It follows that the total number of customers of the H-sector is *Ms*. The customer capacity of the H-sector is instead given by the sum of all workers employed in that sector, times the number of customers a worker can attend, *µ*. Since in equilibrium the sum of all workers employed in the H-sector is the labor supply in the sector, equilibrium customer capacity is given by *µM*(1*−ν_K_*)*ν_H_*. We then assume that in a pre-pandemic equilibrium, the number of shopping trips per person is such that total shopping trips equals customer capacity of the H-sector, that is *s* = *µ*(1 *− ν_K_*)*ν_H_*.

To sum-up, a pre-pandemic equilibrium in sector H is a wage rate *w_H_*, a quantity of H-labor per type of establishment *n_Hj_* and an amount of customers per capital *d*, such that, i) given wages and customers, *n_Hj_* is chosen optimally by the establishment manager, ii) labor market for H-workers clear and iii) the total number of shopping trips equals the customer capacity of the H sector.^6^ Note that our concept of equilibrium guarantees that in every prepandemic period every shopper in each of her/his shopping trip is assigned to an H-worker that can serve her. Note that the maximization of profit at the establishment level, plus the heterogeneity in capital imply that type 2 establishments will employ more labor, make more sales and have higher output.

###### 3.3.2 Production during the pandemic

In the pre-pandemic equilibrium output is equal across establishments of the same type and is constant over time. During the pandemic, however, output can change over time, and it can be different across establishments of the same type. As discussed in Section 3.2 nodes that are infected and show symptoms are prevented from working and shopping. Moreover, as the disease spreads, policies are introduced that may prevent also a fraction of healthy workers from working at their establishment. We denote by *n_Lit_* the number of L-workers that show up at work in establishment *i* in period *t*, by *n_Hjit_* the number for H-workers that show up at work in H-establishment *i* of type *j* (large or small) in period *t*, and finally by *d_it_* the number of customers (per unit of capital) that will show up to shop at H-establishment *i* in period *t*. By assumption, in the short run establishments can not replace workers, therefore when the number of workers falls, establishment output will also fall. Moreover, when a customer assigned to an H-establishment is sick and does not show up to shop, the output of that establishment also is reduced. We can now define *Y_t_*, i.e. the total production of the city in period *t* as

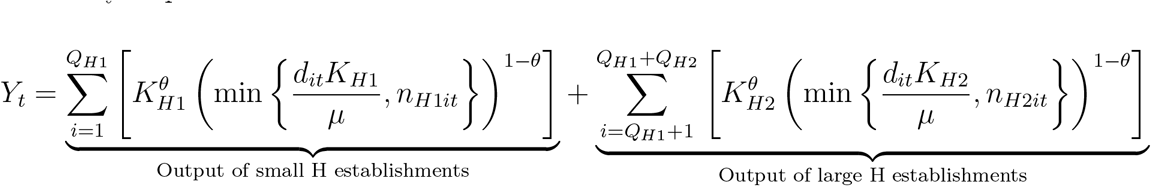

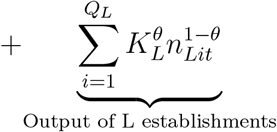

The time series for *Y_t_* during the pandemic is a key object of interest in our policy experiments below, as it summarizes the economic impact of the pandemic and of the various measures of pandemic control.

## 4 Calibration

In this section we describe how we set the values for the parameters of the ECON-EPI network, in order to numerically assess the impact of the pandemic and the effects of several policies.

### 4.1 Demographics and Public Transportation

We calibrate our model to a 5% synthetic version of the New York-Newark-Jersey City (NY-NJ-PA) metro area, which in 2019 had a population of approximately 20 million. The percentage of kids in the population *ν_K_* is set to 28% so that the synthetic city has 40% of households with kids, which matches the percentage of households with kids in the metro area from the 2014–2018 American Community Survey (ACS). The percentage of non working adults *ν_N_* is set to 37%, to match the employment to population ratio for persons over 18 in the metro in 2019.^7^ The share of agents using public transportation, *ϕ*, is set to 32% in order to match the percentage of individuals who report commuting to work using public transportation in the NY-NJ-PA metro area from the 2014–2018 ACS.

### 4.2 Workplace

An important aspect of the calibration is to map workers in the data to workers in the two sectors of the model: the H and L sectors. In order to do so, we first work with occupations. Recall that there are two key features that characterize the H-sector: one is the physical proximity with other people (so that infection can be transmitted) and the second is the instability of the contact with customers (which also speeds up the spread of the disease). To capture these two features in an occupation, we use two questions in the ONET database.

The first one asks about physical proximity to other people on the job, while the second one asks about the importance of interactions with external customers.^8^

The answers to these questions can be used to construct two indexes, both ranging from 0 to 100, that give, for each 6-digit occupation, measures of physical proximity and external interactions. Next, using a standard crosswalk, we compute similar indices for all the private sectors at the 2-Digits NAICS level, where the index for sector *i* is the average of the indices of each occupation *j* in that sector, weighted by the national employment share of occupation *j* in sector *i*. This procedure yields indices of physical proximity and external interactions for all the 2-digits NAICS sector. Figure 4 shows these (standardized) indices for all the NAICS 2 digits private sectors.

**Figure 4:**
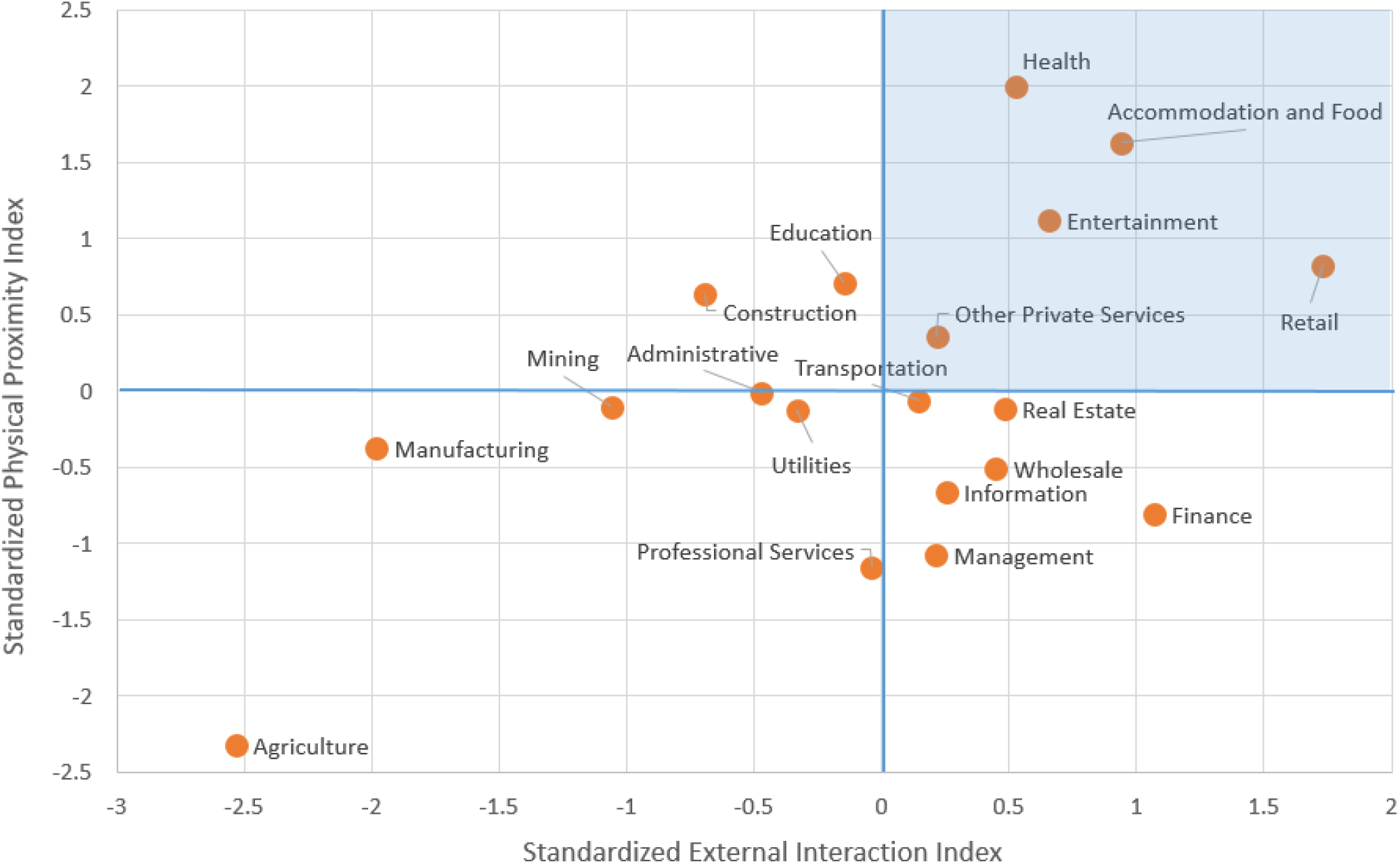
Identifying high contact sectors

The shaded northwest quadrant highlights the 5 sectors which have both indices above the mean; we thus construct the H-sector by aggregating them, and the L-sector by aggregating all the others.

In Table 2, we report key characteristics of workers in the two sectors using employment figures from the Census Statistics of US Business (SUSB) for the NY-NJ-PA metro area in 2016. The L-sector employs more workers (54% v/s 46%), and workers in that sector have higher average yearly wages (94*k* v/s 40*k*). In the last two columns we compute the fraction of workers in each sector that work from home and a measure of their wage premium (relative to those who do not work from home). Note that in the L sector there are many more workers that work from home and that the annual wage of the home L-workers is roughly 16% higher than the wage of the non home workers in the same sector.^9^

In the next section, we use these numbers to pin down the labor supply and the techn-ological differences across the two sectors.

### 4.3 Labor and Technology

The general logic of this section is that restrictions from the pre-pandemic equilibrium (see Section 3.3.1), plus data from firms and workers as described above, pin down the parameters that characterize the labor supply and the technology in the L and H sectors. All the parameters are reported in Table 3 below.

We first use statistics on home workers and on their wages reported in Table 2 to pin down the parameters *ω* and *δ_ω_*, which determine: (i) the fraction of L-workers that can work from home and (ii) the ratio of their wage relative to the wage of those who cannot work from home in the L-sector.^10^ In order to determine the fraction of L-workers who can work from home in our series of experiments, we use two types of information. In Table 2, we report that in the L-sector 7% of workers already work from home before the pandemic hits. However, this is a lower bound for the fraction of workers that can actually start tele-commuting once the level of infections starts to increase and social-distance and lockdown measures are implemented. Dingel and Neiman (2020) estimate the fraction of workers that can potentially work from home in each occupation based on occupational characteristics. We compute their measure for each sector and, aggregating by sector, we find that the fraction of L-workers that can in principle tele-commute is 49.7%. We view this number as an upper bound as, in the short run, it is unlikely that such a large percentage of workers can switch to tele-commuting. For this reason, we set the fraction of workers who can actually work from home once the pandemic hits, *ω*, to 28% (which is the mid-point between the lower and upper bound).

In summary, we consider a pre-pandemic equilibrium with *ω* = 7% and increase the number of L-workers working from home to *ω* = 28% during the pandemic. We assume that non-home workers supply 1 unit of labor input and home workers supply 1+*δ_ω_* = 1.16 units, in order to match the wage differences between the two groups in the L-sector. We then use demographic statistics from Table 1, plus worker statistics from Table 2, to pin down the parameters *ν_L_* and *ν_H_*, which denote the share of adults working in the L and H sectors, respectively. All these parameters determine the total labor supply (in units of labor input) in both sectors.

**Table 1:** Demographics and Public Transportation

| Parameter Name | Symbol | Value | Source |
| --- | --- | --- | --- |
| *Demographics* |  |  |  |
| Total Population | $M$ | 1,000,000 | Census Data: ACS 2018 |
| Share of Kids | $\nu_K$ | 28% | American Community Survey |
| Share of Non-working Adults | $\nu_N$ | 37% | American Community Survey |
| *Public Transportation* |  |  |  |
| Share using Public Transportation | $\phi$ | 32% | American Community Survey |

**Table 2:** Characteristics of workers in H and L Sectors

| | Share | Avg. yearly wages (\$) | Share Home workers | Home wage premium |
| --- | --- | --- | --- | --- |
| L-sector | 54% | 94k | 7% | 16% |
| H-sector | 46% | 40k | 3% | 6% |

**Table 3:** Labor and Technology Parameters

| Parameter Name | Symbol | Value |
| --- | --- | --- |
| Share of Capital Income | $\theta$ | 33% |
| *L-Sector* |  |  |
| Share of adults working in L | $\nu_L$ | 34% |
| Share of L-workers that can work from home | $\omega$ | 28% |
| Home premium | $\delta_\omega$ | 16% |
| Number of establishments (14 wkrs) | $Q_L$ | 18,400 |
| Capital per unit of labor | $\frac{K_L}{n_L}$ | 3.4 |
| *H-sector* |  |  |
| Share of adults working in H | $\nu_H$ | 29% |
| Number of small establishments (4 wkrs) | $Q_{H1}$ | 12,000 |
| Number of large establishments (50 wkrs) | $Q_{H2}$ | 3,500 |
| Number of customers per H-worker | $\mu$ | 10 |
| Capital per unit of labor | $\frac{K_H}{n_H}$ | 0.26 |

Both sectors share constant returns to scale production functions, where capital share is common and given by *θ*. We estimate *θ* using the standard methodology outlined in Cooley and Prescott (1995), using 2018 data for the New York Metro Area.^11^

Given *θ* we can normalize the wage of a unit of labor (which is equivalent to the wage of a non-home worker) in the L-sector to 1 and use the establishment labor demand (Equation 4) to pin down the labor demand per unit of capital. We then use the labor market clearing (Equation 5) to pin down the total capital in the sector *Q_L_K_L_*. Note that in the pre-pandemic equilibrium the number of establishments *Q_L_* is not determined separately from the capital per establishment *K_L_*, so we simply pick *Q_L_* so that the number of workers per establishment in the model is 14, which matches the number of workers per establishment in the NAICS sectors that compose our L-sector.^12^

Now moving to the H-sector, we use, as we did in the L-sector, the establishment labor demand (Equation 6) to pin down the labor demand per unit of capital. We then use the labor market clearing to pin down the total capital in the sector *Q_H_*_1_*K_H_*_1_ + *Q_H_*_2_*K_H_*_2_. We pick *Q_H_*_1_ and *Q_H_*_2_ to match features of the establishment size distribution in the NY-NJPA metro in the NAICS sectors that comprise our H-sector. In particular, we choose the size of the small establishments to match the average establishment size of the firms in the H-sector that have less than 20 employees. This gives a number of 4 employees for the small establishment and 50 employees for the large establishments. This choice, together with equilibrium restrictions, implies that in the model 22% of H-workers are in 4-employee establishments (so that we match the employment share of small establishments). We denote this share as *ν_H_*_1_. The values of *Q_H_*_1_ and *Q_H_*_2_ are reported in Table 3.

The remaining parameter to be determined in the H sector is *µ*, that is the number of customers that a worker can attend to in a day. Recall that, in a pre-pandemic equilibrium, in the H-sector the number of customers is equal to the total customer capacity. In the next section, we calibrate the equilibrium shopping trips (*s*) to be 2 per person, so that the total number of customers in a day is 2*M*. This implies, given the share of H-workers in the population, that the parameter *µ* is approximately 10; that is, an H-sector worker serves an average of 10 customers per day. One final important statistic reported in Table 3 is the capital per unit of labor, which is higher in the L-sector (3.4) than in the H-sector (0.26). The magnitude of this gap is identified from data on the wage differential (see Table 2) between workers in the two sectors. The reason why a unit of labor used in the production of the final good in the L-sector receives a higher compensation than a unit of labor used in the production of the final good in the H-sector, is that labor in the L-sector works with more capital.^13^

### 4.4 Network Contacts and Weights

The number of contacts each person has on each layer, and the weights of different layers play an important role in the spreading of the disease through the network. Our main reference for setting these in the model is the work by Mossong et al. (2008), which, using a common paper-diary methodology, has collected data on various characteristics of daily face-to-face interactions for a sample of over 7000 persons in 8 European countries.

The number of contacts of various individuals in different layers in the model and the targets from Mossong et al. (2008) are reported in the first two columns of Table 4.

**Table 4:** Network Contacts and Weights

| Person type | Layer | Actual Contacts |  | Weight | Contact Pool |
| --- | --- | --- | --- | --- | --- |
|  |  | Model | Mossong |  |  |
| *All:* | Home and Neighbor | 5.2 | 5.2 | [ 22%, 10%] | - |
|  | Shopping overall | 2 | 2 | 10% |  |
| | small | $2v_{H1}$ | | | 4 |
| | large | $2(1 - v_{H1})$ | | | $\tilde{v}v_H M$ |
|  | Public Transport | 0.3 | 0.4 | 4% | 54 |
| *Kids:* | Total | 15.5 | 15.3 |  |  |
|  | School | 8 |  | 22% | 26 |
| *Adults:* | Total | 12.3 | 12.4 |  |  |
|  | Work |  |  |  |  |
|  | H-small (co-workers) | 1 |  | 22% | - |
|  | H-small (customers) | 10 |  | 22% | 56 |
|  | H-large (co-workers) | 4 |  | 22% | - |
| | H large (customers) | 10 | | 22% | $M$ |
|  | L (co-workers) | 3.7 |  | 22% | - |

Mossong et al. (2008) reports that on average each individual has 5.2 contacts in the household and during leisure activities. We map these contacts with model’s contacts that take place within the household and neighborhood layers. Since the average household size in the model is 1.6, we impose that each household has on average 3 neighbors (some households have two neighbors and some have four), so that each individual has an average of 1.6+3*·*1.6*−*1 = 5.2 household/neighbor contacts. Mossong et al. (2008) also reports that, on average, each individual experiences 2 contacts during shopping and 0.4 while traveling. We set the number of shopping trips per person and the number of meetings while using public transportation in the model to match these two figures.

Moving now to the differences between kids and adults, Mossong et al. (2008) reports that kids between the ages of 0 and 19 have on average 15.3 contacts, and adults have on average 12.4 contacts. In the model, we set the number of school contacts (which are specific to kids) to match total kids contacts. For adults, the number of contacts is more heterogenous. A fraction of adults (the non-workers) have no contacts resulting from work. Another fraction (the L-workers) have contacts resulting from meeting their team (of size *T_L_*) of co-workers, where the team of workers is meant to capture the set of co-workers with which a worker interacts more closely. Finally, the H-workers have contacts resulting from the team of co-workers (of sizes *T_H1_* and *T_H2_*) and from meeting with customers (*µ*). Since we do not have much hard evidence on the size of workers teams, we simply set the size of the team of H-workers in the small establishments *T_H1_* to 2 (mom and pop stores) and set *T_H2_* = *T_L_* = 5 so to match the total number of adult contacts.^14^ Notice also that this choice for the size of teams together with the data on establishment sizes in Table 3 implies that an L-establishment employs 3 teams, a H-large establishment employs 10 teams and a H-small establishment employs 2 teams.

Mossong et al. (2008) also reports information on the average duration of contacts, by contact type (daily, weekly and first time). We identify contacts in the household, work and school layers as daily, with an associated average duration of 3 hours. We identify shopping and neighborhood with weekly contacts, with an associated average duration of 1.4 hours and finally we think of contacts during travel as first time contacts, with an average duration of 0.5 hours. These figures results in weights of each layer (normalized to sum to 1) which are reported in the third column of Table 4. These weights are then used to identify the parameters used in equation 2 that capture the weight of each layer *ω^H^* = *ω^W^* = *ω^S^* = 22%, *ω^P^* = 4% and *ω^C^* = *ω^N^* = 10%.

The final column of Table 4 reports the potential pool of contacts for those layers where the actual contacts are drawn randomly every day. This information is not available in Mossong et al. (2008), however it is an important determinant of the spread of infection, and therefore we pin it down using the network structure, as well as additional information. For the shopping links, every person does (on average) 2*v_H_*_1_ and 2(1*−v_H_*_1_) shopping trips to the small and large establishments, respectively. When shopping at the local mom and pop store, the pool of potential sales people that a shopper meets is given by 4 (the employment size of the H-small establishment). When shopping at large establishments, the pool of potential contacts is given by 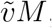, with 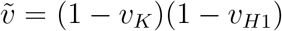, which is the total number of workers that work in large establishments in the H-sector. Note that pool of contacts (i.e. potential sales people) when shopping at large establishments is much larger than the contacts when shopping at the small stores. The reason is the assumption of a different customer base: when shopping at a small establishment, a person always visits the same local store whereas when shopping at a large establishment, the individual is randomly assigned to an establishment in the city.

For adults working in local small establishments in the H-sector, the pool of potential customers is given by the local customer base which is equal to the size of the population divided by the number of workers in the small establishments,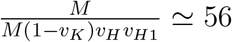. Workers in large establishments in the H sector draw their potential customers from the whole city, so their pool of contacts is the city population *M*.

For public transportation, we choose the number of potential contacts equal to 54 to match the seating capacity of the R160 New York City subway car. Finally, for schools, we proxy the pool of potential contacts with the class, so we set the size of the pool to 26 to match average class size (across grades) in New York City public schools for 2018–19. The ratio between the actual contacts and the contact pool for the unstable layers (shopping, public transportation, school and H-work place) is then used to set the Bernoulli parameter *ρ_i_* in the network clocks described in Equation 1.

### 4.5 Disease Transmission

The final parameters to be determined are those regulating the diffusion of the disease, described in Sub-Section 3.2. We set some parameters based on epidemiological studies on COVID-19, and set the remaining, for which there is less evidence, to match the early stages of infection diffusion in the New York metro. Parameters are reported in Table 5.

**Table 5:** Disease Transmission Parameters

| Parameter Name | Symbol | Value | Target |
| --- | --- | --- | --- |
| Infection Probability | $\pi$ | 0.52 | Calibrated (see text) |
| Relative Infectiousness of $IA$ | $\eta$ | 0.50 | Li et al. (2020) |
| Prob. of transition from $E$ to $IA$ | $\alpha$ | 0.6 | Calibrated (see text) |
| Initial Ratio of Asy. to Sym. | $r_{as}$ | 1.7 | Calibrated (see text) |
| Prob. of transition from $IP$ to $IS$ | $\gamma$ | 0.25 | Incubation 4 days |
| Prob. of transition from $IS$ to $R$ | $\rho_S$ | 0.071 | Duration of disease 14 days |
| Prob. of transition from $IA$ to $R$ | $\rho_A$ | 0.25 | Duration of asymptomatic stage of 4 days |

Starting on the symptomatic branch, we set *γ* to 0.25 and *ρ_S_* to 0.071, in order to match a duration of the pre-symptomatic and symptomatic stages of the disease to 4 and 14 days respectively (see, among others, Guan et al. 2020). Going now to the asymptomatic branch, we follow Li et al. (2020) and set *η* to 0.5, capturing the finding that asymptomatic are half as infectious as the patients showing symptoms. Also, following Li et al. (2020), we set *ρ_A_* to match a duration of the asymptomatic stage to 4 days. The three remaining parameters are *π*, the infectiousness of the symptomatic cases, *α*, the fraction of exposed that transit to the asymptomatic stage, and *r_as_*, the initial ratio of asymptomatic to symptomatic. Our strategy is to pick these parameters so that the infection curve in the model exactly matches the data in the initial period of the infection. In the next section we explain in more detail this choice.

## 5 Results

This section first describes how we use mobility data to discipline changes in the network contacts during the pandemic. It then shows how the calibrated ECON-EPI network performs in explaining the infection dynamics, and contrasts it with another popular model of infection spreading, i.e. the standard random mixing SIR model. Lastly it discusses the contribution of the different layers of the network to the progression of the disease.

### 5.1 Changes in network structure during the pandemic

We focus on the period from March 8th, 2020, where the first 160 cases where reported in the New York metro area, until May 25th, 2020. We start our model city with the same number of infected symptomatic per million in the New York MSA on March 8th. The progress of the infection in the model does not only depend on initial conditions and epidemiological parameters, but also on the network structure which, as the pandemic spreads, evolves. In order to capture this evolution we use both information on actual regulatory changes and data on mobility, as reported by Google. ^15^ In particular Google reports three mobility series that track the visits and length of stay of individuals at workplaces, retail and residences. These series have a natural mapping into our model: workplace mobility maps into presence of L-workers at their establishments, retail mobility maps into presence of workers and shoppers at H-establishments and finally residential mobility captures the time individuals spend at home. These three measures for New York City are reported in the top panel of Figure 5. The panel shows that initially workplace/retail mobility sharply falls, then it stays constant at a depressed level and partially recovers towards the end of the period. Residential mobility displays the opposite pattern. This evolution is most likely the result of both changes in policy and in behavior. Our strategy is to match this evolution by furloughing a time varying fraction of both L and H workers. In particular in each period we match the observed decline in workplace mobility in two ways. In the first days of the pandemic we match the decline in workplace mobility by having all L-workers that can work from home starting to do so. As time progresses and workplace mobility continues to decline we match the further decline by furloughing a fraction of L-workers each day. Then we furlough a fraction of H-workers each day so to match the decline in retail mobility. We impose a larger percentage decline of the employment in large H-establishments, relative to small ones, to be consistent with the fact that authorities in New York shut down events with more than 500 attendees by March 12th.

**Figure 5:**
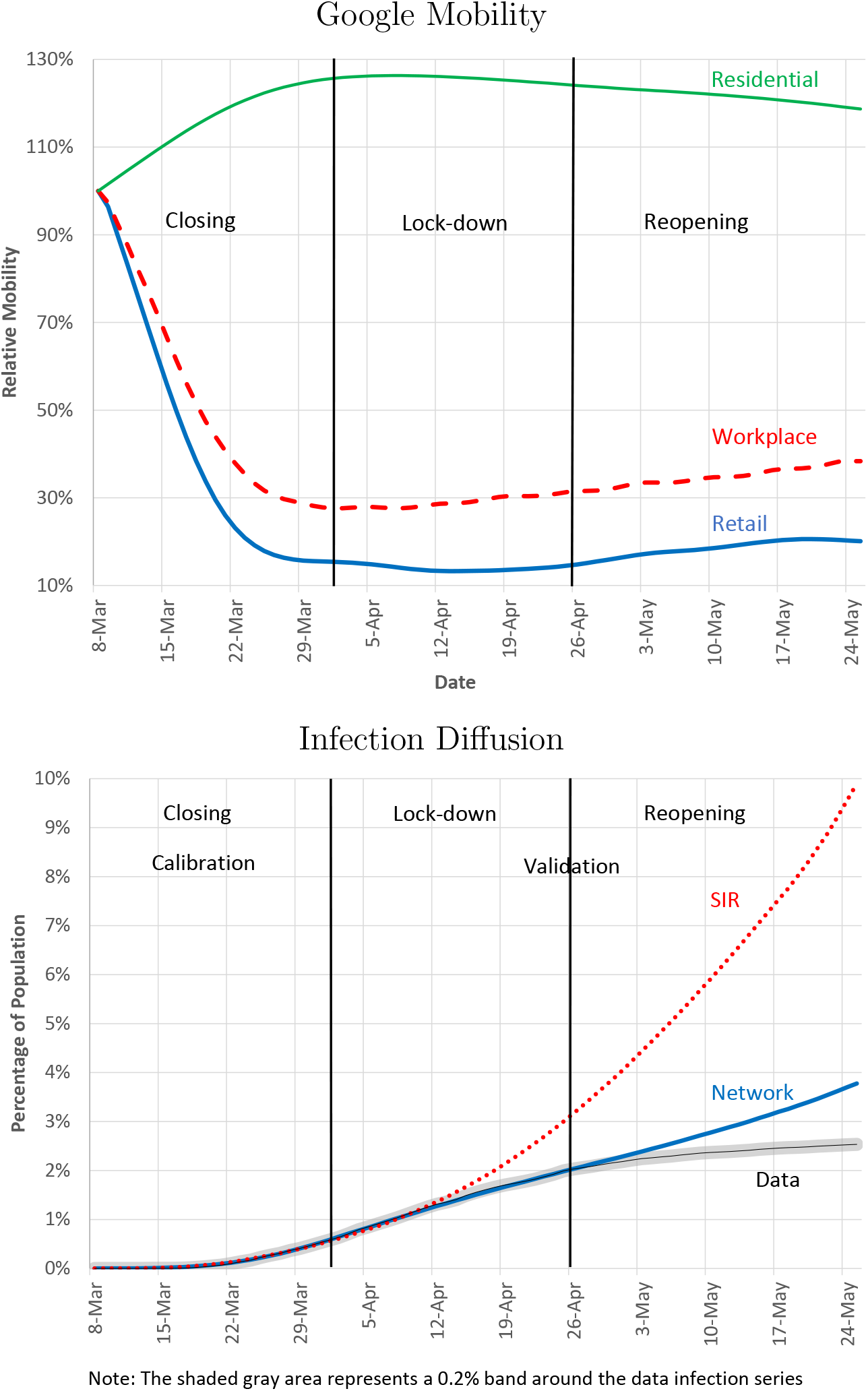
Network v/s SIR

When a worker is furloughed, her time is reallocated to their household and neighborhood networks. A fraction of the work hours are assigned to the household and neighborhood layer so that the increase in home hours matches the increase in residential mobility. Note that when we furlough H-workers we also cut a number of shopping links, as shoppers assigned to furloughed workers are not able to shop. We also close schools in the model on March 14th, which is the date in which K-12 schools are shut-down statewide.^16^

The mobility patterns suggest a division of our period in three subperiods. The first(labeled “closing”, from March 8th to April 3rd) is the period in which mobility sharply declines, the second (labeled “lock-down”, from April 3rd to April 26th) is the period in which mobility stays low, and the last (labeled “re-opening”, from April 26th to May 25th) when mobility picks up.

### 5.2 Network v/s SIR

The bottom panel of Figure 5 shows the cumulative infection curve generated by the network model against the data. Since our calibration strategy is to pick the epidemiological parameters *π, α*, and *r_as_* to match the infection curve in the first sub period, the network model and the data lie on top of each other by construction until April 3rd. The periods after April 3rd, however, constitute a validation of the model. The network model is close to the empirical epidemiological curve all throughout the lock-down phase, and shows more growth in infection (relative to the data) as the city starts to re-open. For comparison purposes, we also report the infection curve predicted by a standard SIR model, where each individual has the same number of contacts as in the network, but the contacts are randomly drawn across the entire population.^17^ We calibrate the epidemiological parameters in the SIR in order to match the data infection curve in the first sub-period (exactly as we did for the network model), and we change the number of random contacts in the SIR so to match the average change in Google mobility. Possibly the most important message of Figure 5 is that even when the two models (Network and SIR) are put on equal footing, as they generate the same initial surge of infection and have similar containment measures, they have sharply different predictions for the evolution of the pandemic. In the network model, the infection naturally slows down, as the reduction in the number of contacts is enough to keep the infection local and prevent the disease from reaching the entire population. The SIR model, however, predicts that despite the reduction in contacts, the infection takes off in an exponential fashion. This is due to the random nature of contacts: in the SIR model, an individual is equally likely to meet any other individual in the city, whereas in the network model contacts are more clustered and less random. Before we move on to policy experiments, we use our calibrated model to quantify the contribution of several layers to the infection.

### 5.3 Infection Decomposition and Complementarities

In this section, we study the effect of shutting down different layers of the network, and how this shutdown interacts with the transmissibility of the disease. In order to do so, we sequentially set to 0 the weights of each layer of the network, and assess the impact of shutting down one single layer on the evolution of the infection. An important issue in assessing the impact of a given layer is the presence of mitigation policies (for example school closures) or endogenous reduction of contacts (as captured by Google mobility). If contacts in a layer are already substantially reduced, we might find that shutting down that layer completely does not have much impact on infection; this, obviously, does not reflect the importance of the layer, but rather the fact that the layer was already almost closed. For this reason, we conduct this experiment in the fully open (pre-pandemic) network.

Figure 6 shows the evolution of the disease under different scenarios. In both panels, we show epidemic curves for the network with all layers open (benchmark), with the large H-establishments shut down, with the L-establishments shut down and finally with schools shut down.^18^ The panel on the left uses the infection probability parameter *π* from our benchmark calibration, while the right panel plots the same curves with a lower infection probability parameter, which we use later in our re-opening experiments.

**Figure 6:**
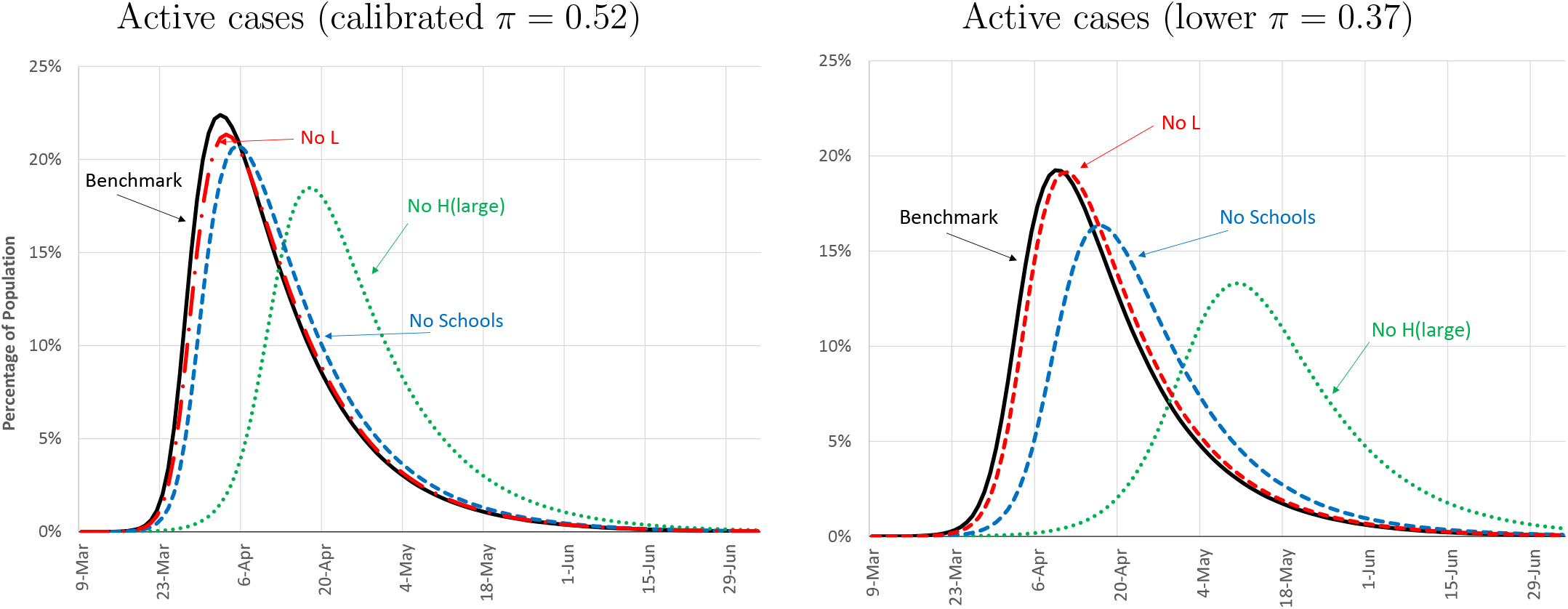
Infection Decomposition

Both panels show that the component of the network which has the biggest impact on infections is the large H-establishments. Shutting those establishments down achieves a substantial delay in the peak of the infection curve and a substantial reduction in the total number of infections. There are two reasons for this. The first one is that, as shown in Table 4, workers in the large establishments in the H-sector have the highest number of contacts, so they are obviously more likely to get sick and spread the infection. The second one is that the customers these workers interact with are randomly drawn from the entire city; this makes their layer very close to a random mixing set-up, and thus very conducive to a rapid spread of the disease. Another layer that is quantitatively relevant is the one related to schools. Table 4 shows that kids in schools have a high number of contacts, also randomly drawn, albeit from a smaller set.

We find it interesting to compare the right and the left panel of Figure 6. The curves in the right panel are drawn from a simulation with a smaller infection probability parameter and thus, not surprisingly, are lower, as there is less infection spreading. Note however that with a smaller *π* the impact of shutting down high contact layers gets magnified. To see why, consider the infection probability of a susceptible node with many infected contacts. If *π* is close to 1, the infection probability is close to 1 and not very sensitive to a marginal reduction of contacts. In this case, shutting down a layer (and thus reducing the number of contacts) does not affect much infection dynamics, which is always very fast. On the other hand, when *π* is lower (but sufficiently far from 0), Equation 3 implies that a marginal reduction of contacts can significantly reduce the infection probability. Therefore, in this case infection dynamics are more sensitive to the network structure, and mitigation policies that reduce the number of contacts are more effective. This highlights an important point, namely the *complementarity* between mitigation policies that reduce the transmission of the disease (e.g. face masks) and mitigation policies that reduce the number of contacts (i.e. shutting down malls). If the transmissibility of the disease is high (*π* close to 1), then a moderate reduction in contacts is not very effective in reducing infections. Similarly, if individuals have a large number of contacts, a moderate reduction in transmissibility is not effective. However, if the transmissibility is lower, then the same reduction of contacts can have a large impact on the spread of the disease, and similarly if the number of contacts is lower, the same reduction in the transmissibility of the disease can have a large impact on infection levels. We will return to these considerations later when we analyze re-opening strategies.

Having established that the network model constitutes a good benchmark to study the evolution of the pandemic, and having analyzed the importance of various layers, we now use the model to conduct two types of policy experiments. The first set, in Section 6.1, studies how counterfactual policies would have affected ECON-EPI outcomes at the outbreak of the COVID-19 pandemic in New York City. These experiments are also helpful to evaluate different options, should a second wave of infections hit. The second set of experiments, in Section 6.2, studies different strategies for reopening the city, as the infection subsides.

## 6 Policy Experiments

### 6.1 Lock-down strategies

As Figure 5 shows, after the draconian lockdown of March and April, infections in the New York metro area stopped increasing by mid May. The question that is often asked is whether the lock-down was too strict. To answer this question, we perform a series of experiments that relax lock-down restrictions in the first four weeks of the pandemic (e.g., between March 8th and April 5th). With the lessons drawn from these experiments, we design a counter-factual *smart* mitigation policy that targets sectors with higher risk of spreading. We show that this policy could have reduced infections and increased output relative to the benchmark case.

We start from our benchmark case and compare it with three counterfactuals in which we gradually bring back the same number of shutdown workers in each sector (L, H-small, and H-large).^19^ We then compare the epidemiological and economic outcomes to our benchmark case. Starting with the epidemiological outcomes (the top panel), we see that increasing workers in the H-large sector has a very large impact (over 1.5% of the population) on infections. Extra workers in the H-small sector have a moderate impact (0.5% of population), while extra workers in the L-sector have almost no impact on the level of infections. The large increase in infections brought about by the additional H-large workers is not surprising; as discussed earlier, these workers have a lot of random contacts, thus they function as spreaders. The sizeable increase in infection coming from bringing back workers in the H-small establishments is more surprising. As discussed in Section 5.3, shutting down these workers when the whole economy is open has no impact on infection dynamics. The reason for this difference is the starting point of the experiment. Adding H-small workers when the economy is substantially shutdown contributes to the spreading, while the marginal contribution of the H-small workers when the economy is fully open is small. Finally, the L-workers constitute highly clustered groups in their respective productive units, who meet frequently and do not interact with customers. For these reasons, a relatively small increase in the number of these workers does not affect infections on the margin.

Moving now to the economic outcomes (the bottom panel), we first observe that the largest output gain (around 2% of GDP) is obtained by adding the H-large workers, followed by an output gain of 1.5% of GDP, obtained by adding L-workers; the smallest output gain (around 1%) is obtained by bringing back the H-small workers. To understand this ranking consider that the marginal productivity of a worker is increasing in the capital-labor ratio of the worker’s sector. In the pre-pandemic equilibrium the capital-labor ratio for the L-workers is higher than the one of H-workers (regardless of the size of the establishment). During the pandemic however, workers in the H-large sector are mostly shutdown, implying that their capital-output ratio is the highest: that explains why bringing them back gives the highest output gain. L-workers and H-small workers are instead shutdown in roughly the same proportion, and therefore, because L-workers have a higher capital to start with, bringing them back results in a larger gain (relative to the H-small).

The results so far suggest that tightening the shutdown in the H-sector while relaxing it in the L-sector might achieve a reduction in infection *and* an increase in output, relative to the benchmark. In Figure 8, we consider the effect of such a policy, which we label smart mitigation. More specifically, we impose stricter lock-down measures in the H-sector (mostly in the H-small sector) while relaxing those in the L-sector. We impose that the total number of individuals going to work is the same as the benchmark and that the amount of workers in each sector affected by the policy is around 1% of the pre-pandemic employment level (the same amount considered in the experiments in Figure 7). A concrete example of such a policy would be to allow more workers in manufacturing plants to go to work, while furloughing an equal number of retail workers that are allowed to go to work in the benchmark. The figure shows that the smart mitigation achieves a substantial double-gain. The top panel shows that it reduces the number of infections by 1.5% of the population of the metro area (300 thousands fewer cases) and the bottom panel shows that at the same time it increases output, relative to the benchmark, by an average of 1%.

**Figure 7:**
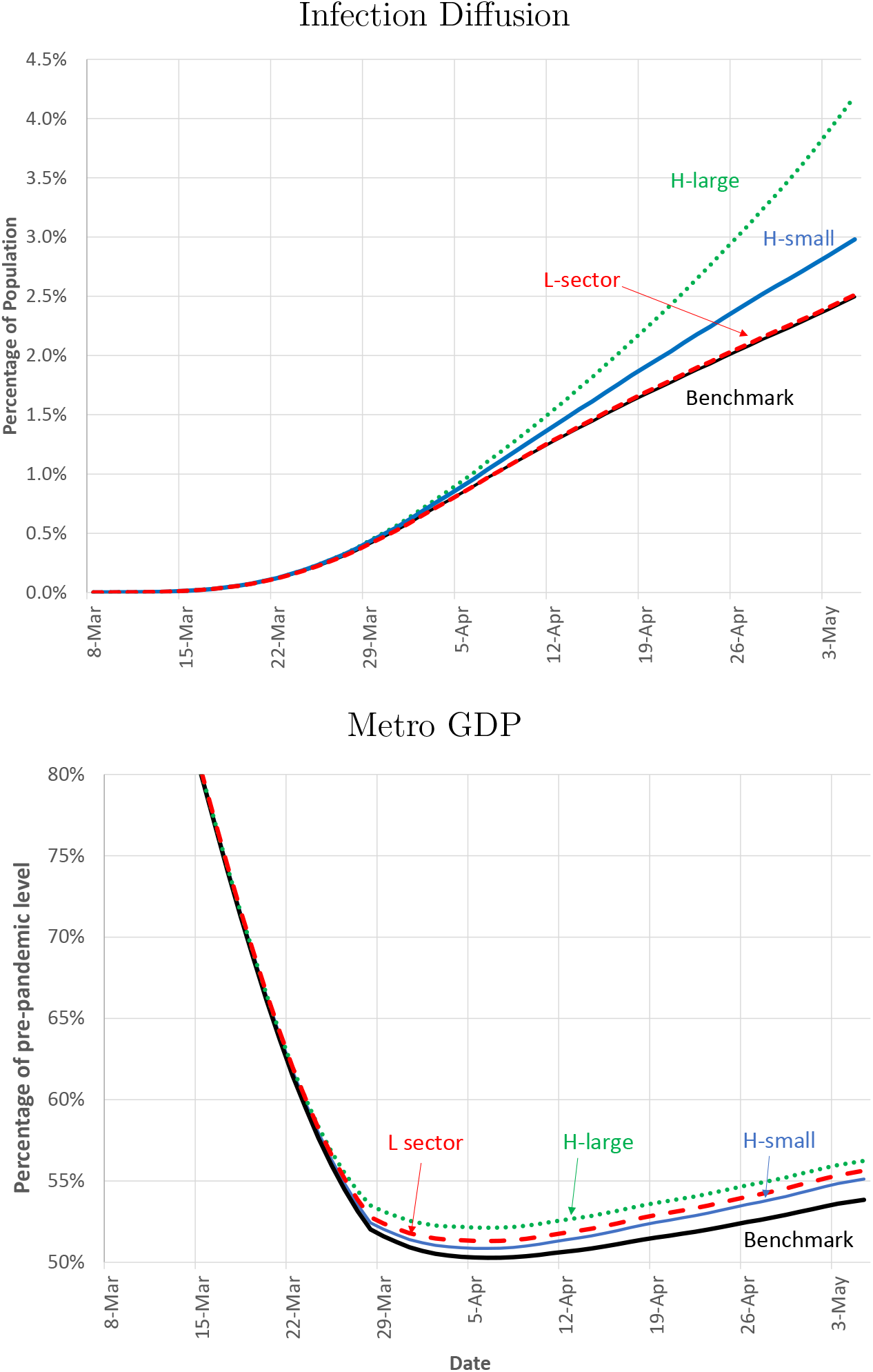
Policy counterfactual

**Figure 8:**
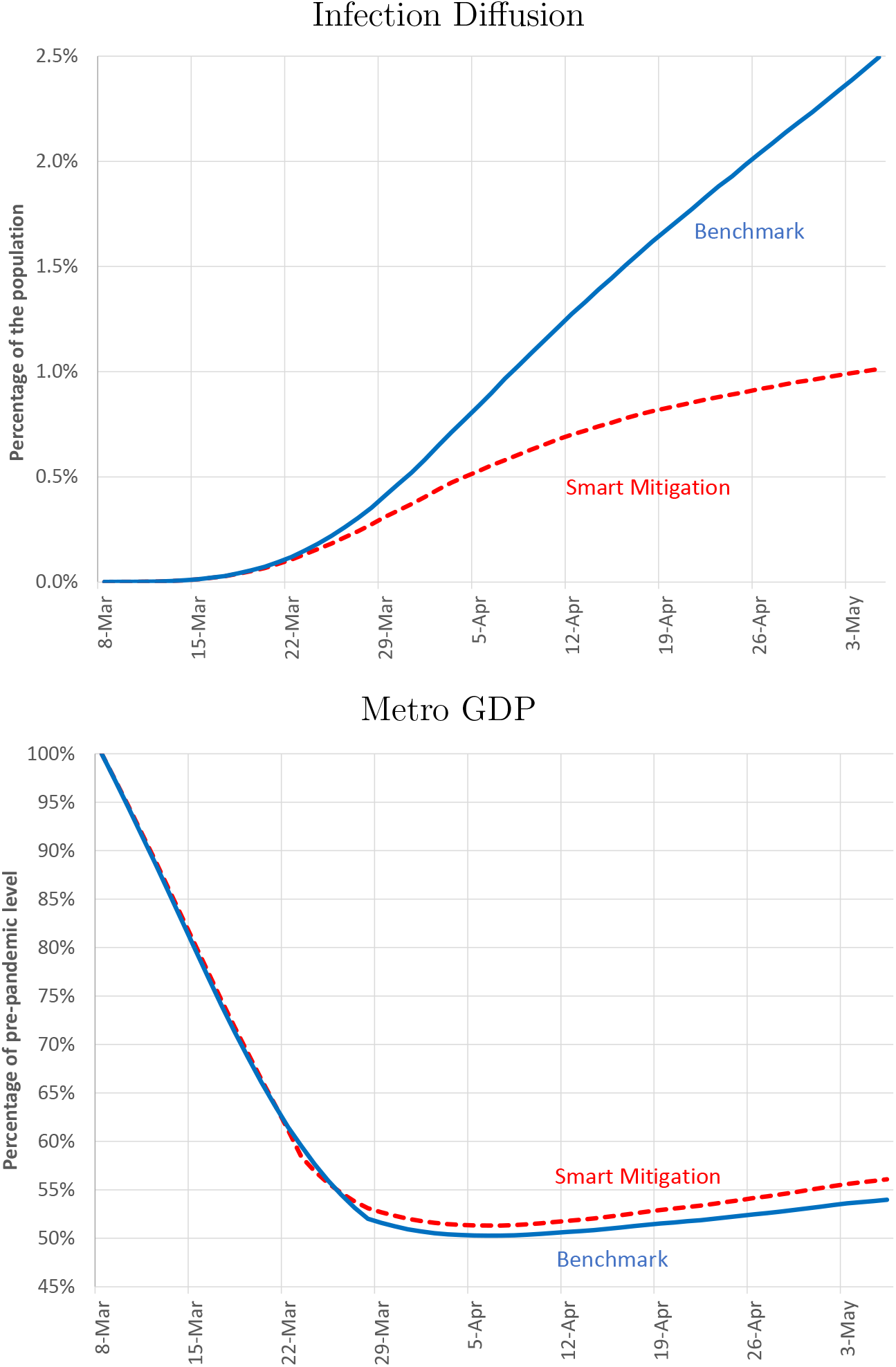
Smart Mitigation

To better understand the source of the double gain one can view this policy as a two steps procedure. The first step is to add workers to the L-sector. As Figure 7 shows, this step involves an increase in output and virtually no change in infection levels. The second step involves a reduction of (mostly) H-small workers. Figure 7 suggests that this causes a substantial reduction in infection and a reduction in output that is smaller than the gain obtained in the first step: hence the double gain.

### 6.2 Re-opening strategies

Results in Section 5 suggest that the network model captures well infection dynamics in the lock-down period. However, as the city starts to reopen in the month of May, the model predicts a level of infection that is higher than the data. One possible reason for this discrepancy is that we keep the epidemiological parameters constant throughout our period, while the much broader availability of PPE and of testing, together with social distancing (e.g. requiring individuals to be 6 feet apart form each other) has reduced the transmissibility of the disease. As this issue is critical to analyze reopening scenarios, we incorporate changes in transmissibility by assuming that in the post lock down period (after April 26th) there is a one-time decline in the parameter *π*. We calibrate this decline (from 0.52 to 0.37,) so that the infection curve in the reopening period (April 26th through May 25th) matches the data. The result of this procedure is illustrated in Figure 9. The figure suggests that the network model with the recalibrated *π* can be a good starting point to study reopening strategies, that is to predict the evolution of infection and output under different assumptions for the evolution of mobility.

**Figure 9:**
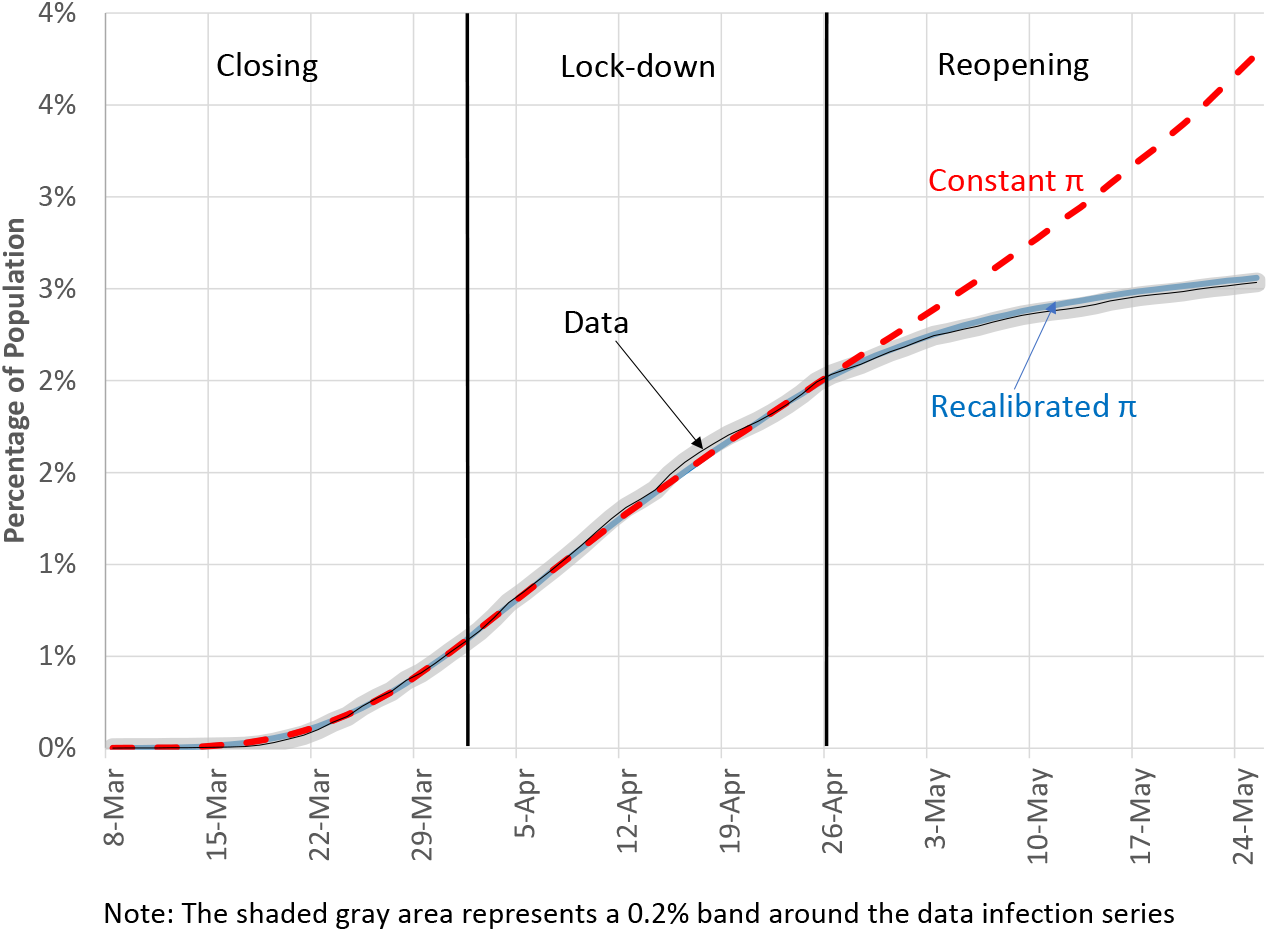
The impact of lower *π*

### 6.3 Re-opening workplaces

In Figure 10 we consider three scenarios for the New York metro. The first (labeled no-reopening) is the one in which mobility stops increasing on May 25th. The second (labeled L-reopening) is the one in which only the L-establishments are allowed to substantially reopen, and the third (labeled H and L reopening) is the one in which both L and large H-establishments are allowed to substantially reopen.^20^ The top panel depicts the infection curves, while the bottom panel shows metro GDP. Under the no-reopening scenario GDP remains severely depressed; on the positive side, the cumulative infection curve becomes flat, suggesting that a prolonged shutdown can stop the growth of the disease and thus eradicate it. The dashed lines show that a substantial reopening of the L-sector comes at virtually no infection cost, and with large GDP gains, as the metro area GDP would recover almost up to 25% of the pre-pandemic level. Finally the dotted lines, displaying the consequences of a reopening of both the L and large H-sector, suggest that this scenario is potentially troublesome. GDP would recover more substantially, but the city would suffer a dramatic second wave, with the total number of infected reaching over 7% of the population by early September. It is doubtful whether in such a scenario the GDP recovery can be sustained.

**Figure 10:**
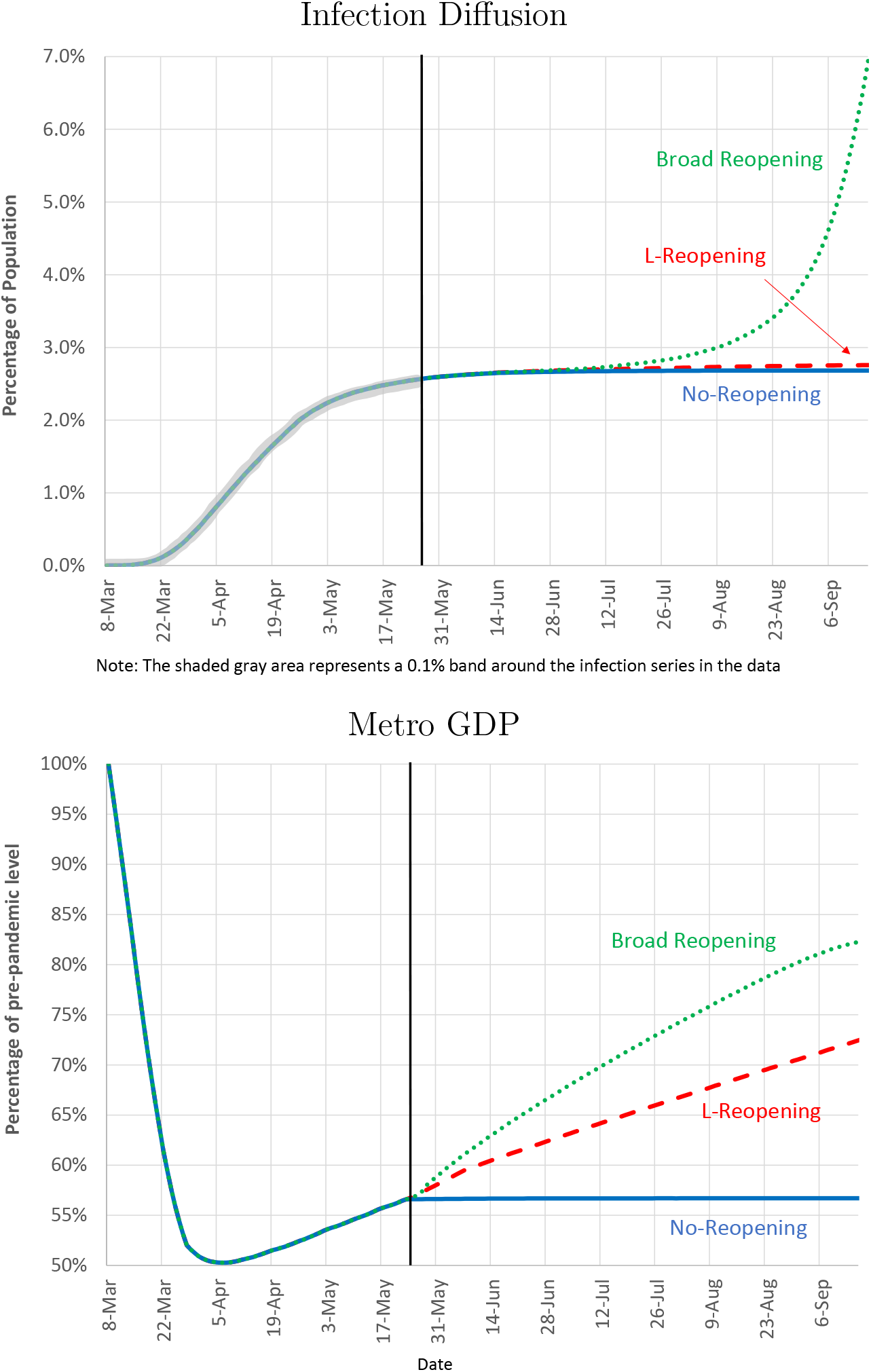
Reopening scenarios

### 6.4 Reopening schools

In the experiments above, we assumed that schools remained closed until the end of the year. One important issue during the COVID-19 pandemic is the impact of reopening activities for the kids, such as schools and summer camps. The impact of reopening these activities on infections depends significantly on the current level of infections, and hence on the date in which the reopening happens. In the top panel of Figure 11, we depict the effects of school reopenings in different dates assuming that mobility in the L and H sectors stays constant at the level of May 25th. On the right scale of the panel, we plot the curve depicting the level of active cases under the scenario of no-reopening of schools. This curve shows that in July, there is still a positive mass of active cases; thus if schools were to re-open in July the large increase in contacts brought about by the open schools would imply a fast growth of symptomatic infections which would reach 10% of the population by mid November. If, on the other hand, schools were to re-open in August, when the mass of active cases is minimal, the addition of contacts from schools would not be causing a rapid takeoff of infections, so the disease would be manageable.

**Figure 11:**
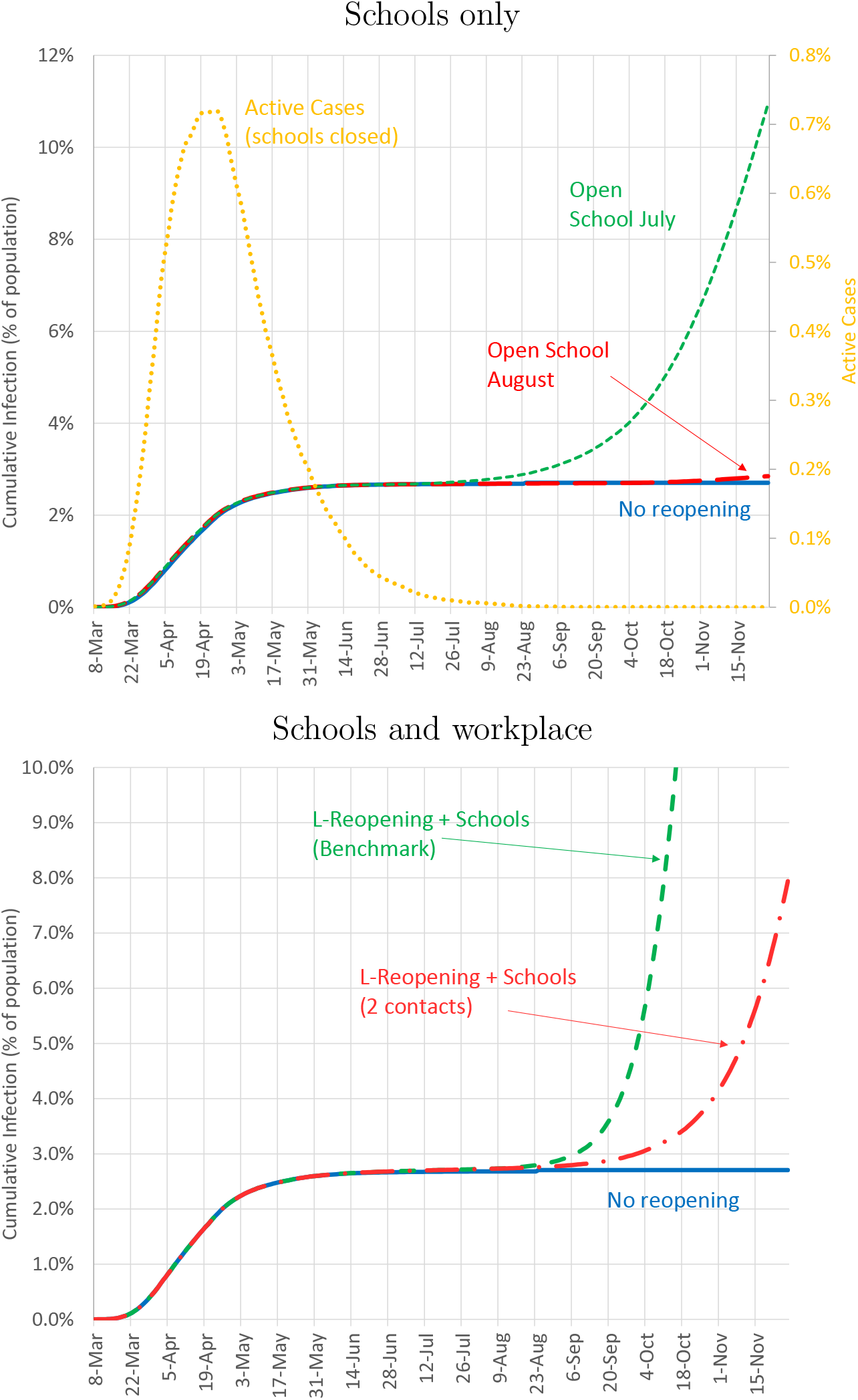
Reopening Schools

**Figure 12:**
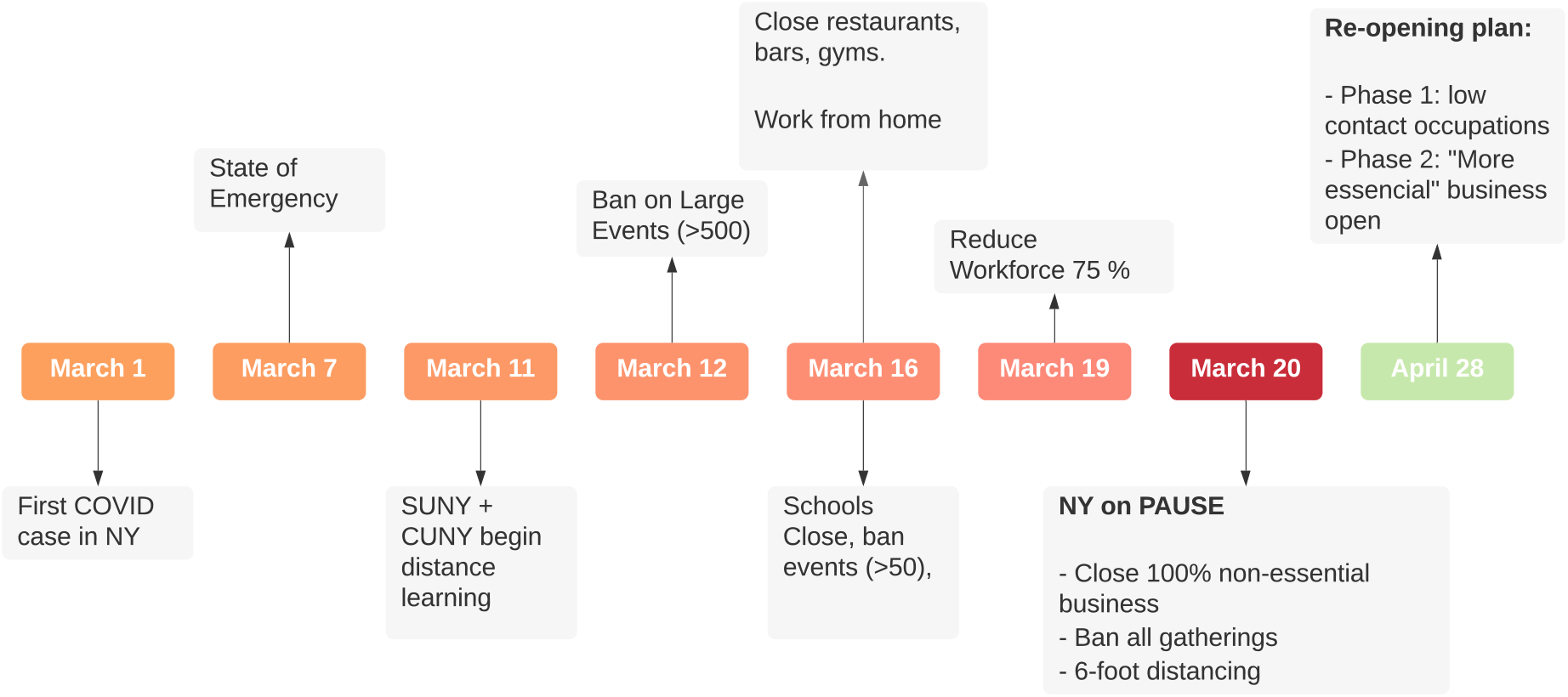
Timeline of lock-down Policies

In our final experiment, we consider reopening scenarios that combine increases in mobility of the L-sector (such as those in Figure 10) with school reopenings. We consider two scenarios: the benchmark case in which schools reopen with the normal number of contacts, and a socially distanced scenario where contacts are reduced to 2 per kid. We assume that schools open on August 1st, and that school-related activities end by Thanksgiving day. The progression of infections is shown in the bottom panel of Figure 11. The key result is that, even in the case of strongly socially distanced schools, the infection takes off rapidly and exceeds 5% of the population by November. This happens because the impact of reopening the L-sector, despite being fairly modest in itself, is sufficient to keep the number of active cases high enough, so that when schools re-open in August (and the L-sector remains open) infections take off. This experiment highlights the importance of interactions among social layers in our network. Reopening just the L-sector or just schools (in August) results in a relatively small increase in infections, but opening both simultaneously results in a distinctive second wave during the Fall. These experiments also confirm the point in Section 5.3 about the importance of limiting contacts even when the transmissibility of the disease is lower. All the reopening exercises are done with a lower transmissibility parameter *π*, and they all show that a relatively small change in the number of contacts can change the pattern of the disease from eradication to fast diffusion. Besides the complementarity in infection there could be another important complementarity between schools and work, since, when schools are closed, parents have reduced ability to go to work. So far we have abstracted from this issue, but we conjecture that modeling it explicitly would make a stronger case for an early shutdown that would allow schools to re-open.^21^

## 7 Conclusion

We develop an ECON-EPI network model to study the impact of the COVID-19 pandemic on a large US metro area, and to evaluate policies that limit the human as well as the economic damage. We build on the traditional SIR model by using network theory to put structure on the patterns of human interactions. We find that this structure is useful to understand observed epidemiological curves, featuring a large initial surge and a plateau at a relatively low level of infections. Moreover we use our set-up to quantify how layers of interactions contribute both to infection levels and economic activity. Network layers that feature numerous and unstable contacts (such as large gatherings or schools) work as ignition rods for the infection. Smart lock-down policies shut down these layers early, and smart reopenings keep them closed for longer. Opening sectors where workers interact with each other in stable teams (such as manufacturing) is the best strategy to minimize output losses, while at the same time keeping the spread of the disease under control.

There are several directions in which we could expand the study of pandemic control on ECON-EPI networks. In our framework interactions are, for the most part, exogenously determined. One direction for further research would be to study how the ECON-EPI pattern of contacts can change endogenously, both in the short run, in response to fear, and in the long run, in response to increased risk of a new pandemic.^22^

Our network analysis can also prove useful to think about how to efficiently allocate limited testing resources. The same principles we used to design “smart” lockdown and reopening policies, can be used to design “smart” testing. We conjecture that it would be efficient to allocate testing to layers of the network which have more numerous and more unstable contacts, and our framework could be used to quantify the effects of such a policy.^23^ Another extension of our analysis would be to introduce more group level heterogeneity, such as different communities/neighborhoods in the city. Such an extension would help to understand how much of the observed large differences in disease outcomes across groups can be explained by differences in their social structure.^24^ It could also help to design social policies that protect the more exposed communities and, at the same time, reduce average spread. Finally, a related application of our analysis would be to analyze how much of the differences in epidemiological and economic outcomes across metro areas and across countries can be explained by differences in the network of interactions.

## Data Availability

No data availability

## A Changes in regulation in the New York Metro

The sequence of measures imposed by the NY government, aimed at slowing down the spread of the disease, is summarized in Figure 12. Increasingly stricter mitigation policies reducing gatherings, retail and production activities were implemented in a short span of time.

## JEL Classification

D85, E23, E65, I18

## Footnotes

* We thank Maria Cristina De Nardi, as well participants at several seminars and conferences for great comments and suggestions. Also many thanks to Dhananjay Ghei and Thomas Gill for outstanding research assistance. The views expressed herein are those of the authors and not necessarily those of the Federal Reserve Bank of Minneapolis or the Federal Reserve System and CEPR

1 See Keeling and Eames (2005) and Jackson (2010) for excellent surveys of the literature on networks in epidemiology.

2 This heterogeneity has been explored in Acemoglu et al. (2013), Acemoglu et al. (2010) and Azzimonti and Fernandes (2018) in information networks.

3 See Harris 2020 for a study on the role of public transportation in spreading the COVID-19 pandemic in New York City

4 See, for example, Li et al. 2020.

5 The details about mapping actual sectors of the economy into these two stylized sectors are discussed in Section 4.

6 For simplicity, we do not develop an explicit theory of the individual choice of shopping trips. A possible way of doing so, that would be consistent with our equilibrium restriction, would be to have the individual benefit of shopping trips to be decreasing in the tightness of the shopping market, i.e. in the ratio between shoppers and customer capacity

7 Employment figures are from the BLS and population figures are from the Census.

8 Specifically the first question (ONET question 21) is “How physically close to other people are you when you perform your current job?” and the second question (ONET question 8) is “In your current job, how important are interactions that require you to deal with external customers (as in retail sales) or the public in general (as in police work)?”

9 To measure the share of workers that work from home we first use ACS data to compute the share of home workers in each 2-digits NAICS sector and then take a weighted average of these percentages, where the weights are the employment shares of each NAICS sector in our 2 macro sectors. Similarly to compute wages of home workers we take a weighted average of the wages in each sector, where the weights are the shares of home workers in each NAICS sector.

10 Recall that, since few workers in the H-sector work from home, we assume that the percentage of H-workers that work from home is 0, both before and during the pandemic.

11 The estimate is the ratio between capital income (consumption of fixed capital plus rent, interest and dividend income) and the sum of capital income plus labor income (compensation of employees). Data for rent, interest and dividend income, and for compensation of employees is available from BEA regional tables. Consumption of fixed capital is computed by first taking the ratio of consumption of fixed capital to GDP on national data for 2018 (the ratio is 16%) and then multiplying it by the metro area GDP.

12 In the model, the number of workers per establishment is smaller than the quantity of effective labor as the average worker, due to higher productivity of home workers, supplies more than one unit of effective labor.

13 In our set-up, we have abstracted from differences in human capital among the workers in the two sectors, and attributed all the differences in wages to differences in physical capital. Since in the short run physical capital is fixed, the results concerning output losses from shutting down workers in the two sectors are independent on whether we attribute wage differences to physical or human capital.

14 A team size of 5 will result in 4 co-worker contacts for the H-worker, and only 3.7 contacts for the L-worker because a fraction of the workers work from home.

15 See appendix A for a timeline of the pandemic related policies in New York.

16 Schools were announced to be closed on March 16th, a Monday, so we shut down schools effectively on Friday 14th.

17 We do not directly use the SIR model, but an equivalent network formulation. Rather than have multiple network layers, each individual has a single layer which connects them to all other nodes. The transition between health states is regulated by the same parameters as in the network model, and described in Figure 3. The probability of infection is therefore determined by the epidemiological parameters *π* and *η* and the per-period number of contacts. The pre-pandemic number of contacts is set to the average number of contacts across children and adults reported in Mossong et al. (2008), and each period this number is adjusted to match the average change in the Google mobility reports. The parameters *π, r_as_*, and *α* are then calibrated to match the early stages of the pandemic, and take on values 0.48, 1.69 and 0.63, respectively.

18 We do not plot curves for public transportation, and small H-establishments as they have a very small impact.

19 The increase in the amount of workers in each sector is around 1% of the pre-pandemic total employment.

20 Across the three experiments the reopening pattern of the small H-establishments is kept constant, and schools are kept closed.

21 For an interesting analysis of the effect of school closing on work choices of men and women see Alon et al. (2020).

22 See the recent literature on the COVID-19 pandemic studying behavioral responses to the infection, such as Alfaro et al. (2020), Farboodi et al. (2020), Krueger et al. (2020) and Toxvaerd (2020). See Fogli and Veldkamp (2020) for a study of the endogenous evolution of network of interaction in societies with difference prevalence of diseases.

23 For some early works on efficient testing using the standard SIR set-up see Berger et al. (2020) and Chari et al. (2020).

24 For evidence of local differences in disease outcomes in the New York metro see Almagro and Orane-Hutchinson (2020).

